# Community Care of Eating Disorders: Comparison of Online and In-Person Treatment – A Systematic Review and Meta-Analysis

**DOI:** 10.64898/2026.11.11.26367167

**Authors:** Nilima Hamid, Paul Robinson

## Abstract

**Objective:** Eating disorders are serious conditions which sometimes require intensive treatment and are undertreated worldwide. Care can be provided in the community in different treatment delivery modes that could increase access to evidence-based treatment and avoid the negative impacts of delayed interventions. This systematic review and meta-analysis investigated the comparative effectiveness of adult eating disorder community care delivered via in-person, online and mixed (blended/hybrid) modes to provide a global perspective and recommendations for the use of telemedicine in the field.

**Method:** The databases MEDLINE, Embase, PsychINFO, Emcare, Scopus and Web of Science were searched on 22^nd^ June 2025. Inclusion criteria specified quasi-experimental studies which compared two or more treatment delivery modes. The Joanna Briggs Institute risk of bias tool for quasi-experimental studies was used for quality assessments. Qualitative synthesis and meta-analyses were conducted.

**Results:** The daycare/intensive outpatient programme studies showed non-inferiority of in-person and online treatment programmes: EDE-Q Global score (−0.00 [95% CI: −0.23, 0.22]); anxiety (GAD-7/OASIS) (−0.05 [95% CI: −0.31, 0.20]); BMI (0.06 [95% CI: −0.20, 0.33]); and depression (BDI-2/PHQ-9/CES-D) (0.07 [95% CI: −0.31, 0.44]).

The outpatient studies showed non-significant differences between in-person and online treatment programmes: anxiety (GAD-7/PROMIS) (0.28 [95% CI: −0.48, 1.04]); depression (PHQ-9/PROMIS) (0.58 [95% CI: −0.59, 1.75]); and EDE-Q Global score (0.73 [95% CI: - 0.91, 2.36]). For all measures, better outcomes were indicated for online care.

**Discussion:** Comparisons with mixed treatment delivery mode are lacking, but findings provide credence to the viability of online care as being on par or even better under certain conditions than in-person care.

**Summary:**

- Daycare/intensive outpatient programmes for adults were non-inferior delivered in-person or online for BMI, eating disorder, depression, and anxiety symptoms.
- Outpatient programmes for adults showed non-significant differences delivered in-person or online for eating disorder, depression, and anxiety symptoms. But findings suggest a possible superiority of online treatment.
- Research on community care delivered in mixed mode is sparse.
- The evidence base is very small, particularly for the outpatient studies indicating the need for more high-quality quasi-experimental studies and ideally well designed randomised controlled trials.

## INTRODUCTION

Eating disorders (EDs) are serious conditions which sometimes require intensive treatment and are undertreated worldwide (Fairburn & Patel, 2014). Care can be provided in the community (Abbate-Daga et al., 2015) in different treatment delivery modes that could increase access to evidence-based treatment, at lower cost and avoid the negative impacts of delayed interventions, such as hospitalisation and chronic illness. Due to the severe consequences of untreated EDs, alongside promotion of understanding about EDs, prevention and early detection, the prompt and effective treatment of EDs is imperative. In the absence of randomised controlled trials (RCTs) the next best evidence base quasi-experimental (QE) studies were used in this systematic review and meta-analysis to investigate the comparative effectiveness of in-person, online and mixed (blended/hybrid) ED community care for adults to provide a global perspective and recommendations for the use of telemedicine in the field.

Telemedicine is particularly important in unprecedented times, such as the COVID-19 pandemic, when in-person treatment is not possible. Although improvement may take longer than in in-person treatment (Baudinet et al., 2023; Zimmerman et al., 2021), virtual ED treatment showed promise as an effective, accessible and scalable treatment option prior to the COVID-19 pandemic especially in increasing treatment capacity and addressing barriers to treatment such as stigma, time constraints, lack of local services, long waiting lists (Hamid, 2023) with the caveat that in some circumstances telemedicine may have detrimental impacts. For example, age and race disparities may be exacerbated (Rivera et al., 2021), it may be difficult to separate the home and treatment environment as well as increased social isolation (Catenacci & Couturier, 2023). Group therapy can be difficult to implement online due to lack of connection between patient and therapist (Schlissel et al., 2023). Further problems of virtual ED treatment include Internet connectivity issues, reduced therapeutic alliance (Shaw et al., 2021), practical difficulties of observing online meals (Brothwood et al., 2021), anxiety about seeing self on video, and the increased opportunity for patients to be deceptive and mask their symptoms (Fernandez-Aranda, Casas, et al., 2020). However, while online ED treatment proffers less accountability, it may reduce the competitive learning of harmful ED behaviours between patients that can occur in in-person treatment (Feldhege et al., 2021; Zipfel et al., 2002); there is increased parental involvement, patients can have their culturally appropriate home food (Catenacci & Couturier, 2023) and despite emotional aspects and creative group sessions in particular being harder to address, practical skills such as problem-solving may be easier to implement online (Baudinet et al., 2023).

The COVID-19 pandemic caused increased ED symptoms and increased rates of presentation and referrals for EDs of all types across all age groups to both inpatient and community medical settings (Devoe et al., 2023; Friars et al., 2023). In a systematic review it was found that compared to pre-pandemic, there was a 48% increase in hospital admissions across different countries during the pandemic (Devoe et al., 2023). Barrett and Richardson (2020) and Campbell et al. (2022) found that the pandemic resulted in ED referrals being made at lower BMIs, shorter illness duration, and higher levels of medical compromise. Silen and Keski-Rahkonen (2022) observed that the increased incidence and hospitalisation rates for EDs during the pandemic were a consequence of the restrictions imposed on daily activities including exercise, lack of social support and isolation, harmful social media exposure, and increased difficulties in accessing treatment. Virtual treatment allows continuity of care when in-person care is not possible. Due to the increased pressure on ED services and the restrictions imposed, the COVID-19 pandemic caused a rapid shift from in-person consultations to online and mixed community treatment delivery (Plumley et al., 2021). Virtual ED treatment has continued into the post-pandemic era and is proliferating. While this transition has been largely successful for outpatient ED care, daycare (DC) presents more challenges (Gorrell et al., 2022), and research is needed to understand how to exploit these novel modes of treatment delivery to optimise eating disorder treatment.

This systematic review and meta-analysis aimed to undertake an exhaustive search of the literature to identify all quasi-experimental (QE) studies reporting the comparative effectiveness of in-person vs. online vs. mixed (blended/hybrid) ED community care for adults. It is contended that this systematic review and meta-analysis is the first to synthesise all the research comparing the effectiveness of different modes of ED community care delivery for adults globally.

## METHOD

This systematic review and meta-analysis was conducted as part of the thesis submission for the award of PhD in Medicine (University College London, UK) and in accordance with the Preferred Reporting Items for Systematic Reviews and Meta-Analyses (PRISMA) guidelines (Moher et al., 2009; Page et al., 2021). The search, screening, quality assessment and data extraction processes were undertaken independently, with PhD supervision. The study aimed to investigate the comparative effectiveness of in-person vs. online vs. mixed (blended/hybrid) ED community care delivery in adult services globally.

### Eligibility Criteria

Using the PICOS framework (Moher et al., 2009) inclusion and exclusion criteria were specified as:

***Population:*** Adults (18+ years) with any ED treated in adult ED services in any location globally. It is important to note when most participants were over 16 years of age, these studies were included in this systematic review as per the Cochrane guidance (Higgins & Thomas, 2019, p. 36). ***Intervention:*** any mode of ED community care (in-person, online or mixed) except for intensive home treatment or primary care. ***Comparator:*** any mode of ED community care delivered differently (in-person, online or mixed) except for intensive home treatment or primary care. ***Outcome:*** Patient reported outcome measures (PROMs) - change in BMI, improvement in ED symptoms, comorbidities and psychological wellbeing. ***Study Design:*** Quasi-experimental studies (Q-E, naturalistic studies/natural experiments) of between group comparisons (interventions)

There were no date, geographical, gender or language restrictions, but only English language reports were identified. To optimise the evidence included, only studies published in peer-reviewed journals were considered; not grey literature such as study protocols or conference abstracts.

### Search Strategy

The electronic databases MEDLINE, Embase, PsychINFO, Emcare, Scopus and Web of Science were searched from inception through 22^nd^ June 2025. No further articles were identified by database alerts and an updated search on 15^th^ October 2026. Forward and backward citation searching in eligible articles were used to identify further reports. The search strategy for OVID MEDLINE is shown in Table 1.

**Table 1.**
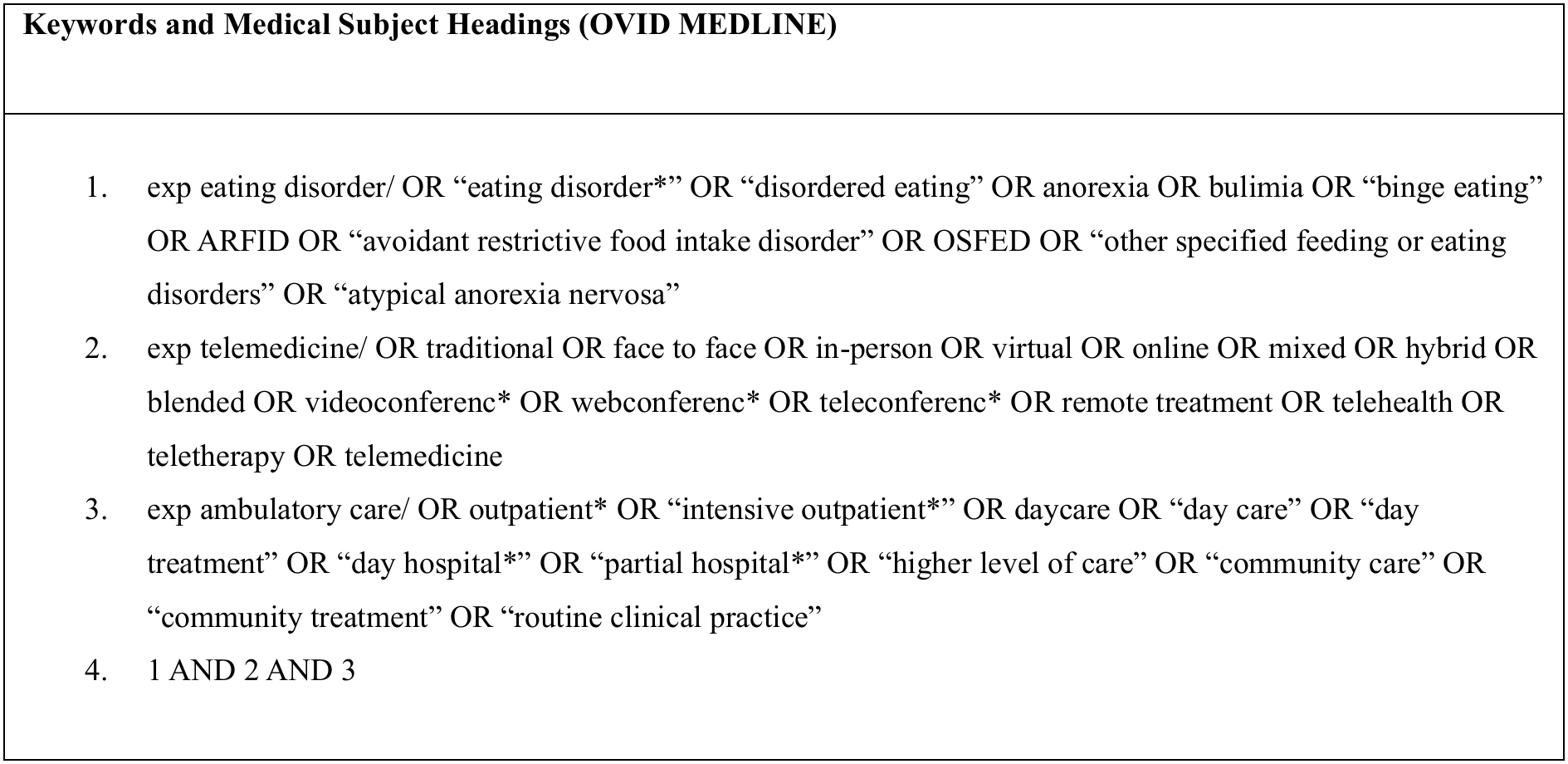
Search Strategy for Databases.

### Quality Assessments

The risk of bias in the included studies was critically appraised using the Joanna Briggs Institute (JBI) tool for critically appraising quasi-experimental (Q-E) intervention studies. The JBI tool for Q-E studies assesses whether precautions were used to mitigate risk of bias and to manage other factors linked to study quality. The questions are answered as being met (yes), unmet (no), unclear, or not applicable; and assessed at study, outcome or result level as appropriate. This tool only considers the internal validity (biases due to study design) and statistical conclusion validity (biases due to methods of statistical analysis) constructs. A qualitative judgement is then made to rate the overall bias in a study as low, moderate, high, or critical risk, but this judgement excludes the statistical conclusion validity construct as only the internal validity construct affects risk of bias (Barker et al., 2024).

### Data Extraction and Analysis

Study and participants’ characteristics, study findings, as well as means and standard deviations for selected outcomes and sample sizes from the included studies were extracted into Microsoft Word tables. Following narrative synthesis, random effects meta-analyses were conducted using IBM SPSS Statistics (Version 30) and a change score calculator generated using Claude AI (https://claude.ai accessed 7 July 2025).

Change-from-baseline between group effect size, standardised mean difference (SMD) was the effect size used in the daycare/intensive outpatient programme (DC-IOP) meta-analyses. Post-treatment between group effect size (SMD) without using change scores was used in the outpatient treatment (OP) meta-analyses. The SMDs, in this case Hedges’ (adjusted) *g* are interpreted according to the thresholds provided by Cohen (1988) of 0.2 = small effect size, 0.5 = medium effect size, and 0.8 = large effect size. It was planned that in the event of insufficient data post-treatment between group meta-analyses (without using change scores) would be conducted.

An inverse variance, random effects model was used and is the default choice for meta-analysing Q-E studies as heterogeneity is expected to be high. The extent of heterogeneity, I^2^ (0% to 100%) was calculated. I^2^ thresholds are low at < 40%; moderate at 30-60%; substantial at 50-90%; and considerable at 75-100% (Higgins & Thomas, 2019, p. 259).

To assess non-inferiority of the intervention (in-person) and comparison (online) groups, the non-inferiority margin was set at Hedges’ *g* < 0.1. Thus, an effect size *g* < 0.1 was considered acceptable for demonstrating non-inferiority, ideally with the whole 95% confidence interval within the region *g* = +/−0.1 (Irish et al., 2022).

The strength of the whole body of evidence was assessed using the Grades of Recommendation, Assessment, Development and Evaluation (GRADE) approach which is based on five criteria: indirectness; study limitations - risk of bias; inconsistency; imprecision; and publication bias (Higgins & Thomas, 2019, p. 389). Confidence in the effect estimates can be categorised as ‘high,’ ‘moderate,’ ‘low’ or ‘very low.’ Non-randomised studies of interventions (e.g. Q-E studies) start with “low” confidence in the effect estimates because of the potential bias from lack of randomisation (i.e. selection bias and confounding) (Higgins & Thomas, 2019, p. 391). The GRADE approach allows Q-E studies (observational) to be upgraded according to three criteria: large effects, dose-response and plausible confounding (Higgins & Thomas, 2019, p. 396).

Various standardised, reliable, and validated instruments which are established in the ED field were consistently used in the included studies. The outcome measures selected for meta-analysis are shown in Table 2.

**Table 2.** Outcome Measures Meta-Analysed.

| **Outcome Measure** | **Description** |
|---|---|
| *Eating Disorder Examination Questionnaire (EDE-Q) (Fairburn & Beglin, 1994)* | Measures eating disorder cognitive (attitudinal) psychopathology on four subscales (dietary restraint and eating, weight, shape concerns) and a global score with items rated for the past 28 days. Eating disorder (binge eating and compensatory) behaviours are rated as the number of episodes in the past 28 days but do not contribute to the subscale scores. Objective bulimic episodes (OBEs) are differentiated from subjective bulimic episodes (SBEs). |
| *Beck Depression Inventory (BDI-2) (Beck et al., 1988)* | A 21-item screening tool that assesses the intensity of depression as minimal, mild, moderate or severe rated for the past two weeks. |
| *Centre for Epidemiological Studies Depression Scale (CES-D) (Orme et al., 1986)* | A 20-item measure determining severity of depression rated for the past week. |
| *Patient Health Questionnaire (PHQ-9) (Kroenke et al., 2001)* | A brief 9-item measure that assesses the severity of depression as mild, moderate, moderately severe or severe rated for the past two weeks. |
| *Generalized Anxiety Disorder (GAD-7) (Spitzer et al., 2006)* | A brief 7-item screening tool that assesses the severity of generalised anxiety disorder rated for the past two weeks. |
| *Overall Anxiety Severity and Impairment Scale (OASIS) (Norman et al., 2006)* | A brief 5-item measure that can be used to assess severity across anxiety disorders and subsyndromal anxiety rated for the past week. |
| *Patient-Reported Outcomes Measurement Information System (PROMIS) – Anxiety Scale – Short Form 8a (Pilkonis et al., 2011)* | A brief 8-item measure assessing severity of anxiety rated for the past 7 days |
| *Patient-Reported Outcomes Measurement Information System (PROMIS) – Depression Scale – Short Form 8a (Pilkonis et al., 2011)* | A brief 8-item measure assessing severity of depression rated for the past 7 days. |
| *Body Mass Index (BMI)* | A measure of body fat based on height and weight. Increased admission to discharge BMI indicates a positive outcome - weight gain from treatment. |

None of the meta-analyses included a minimum of 10 studies, thus no funnel plots were undertaken (Higgins & Thomas, 2019, p. 365). Instead, publication bias was explored by comparing fixed and random effects estimates and visually inspecting forest plots for trends indicating possible publication bias (Higgins & Thomas, 2019, p. 367). Sensitivity-analyses were performed to evaluate the robustness of the results to variations in quality of the studies, type of ED, and to explore heterogeneity.

## RESULTS

In all 2193 reports were identified, 891 abstracts were screened, and 11 full text reports were assessed for eligibility. Nine single report quasi-experimental (Q-E) studies were included in this systematic review and meta-analysis. However, as full data for Robinson et al. (2024) were not available, and there was insufficient information to assess study quality this study is described but not critically appraised or meta-analysed (N = 978: 8 studies). The study selection process is described in Figure 1.

**FIGURE 1.**
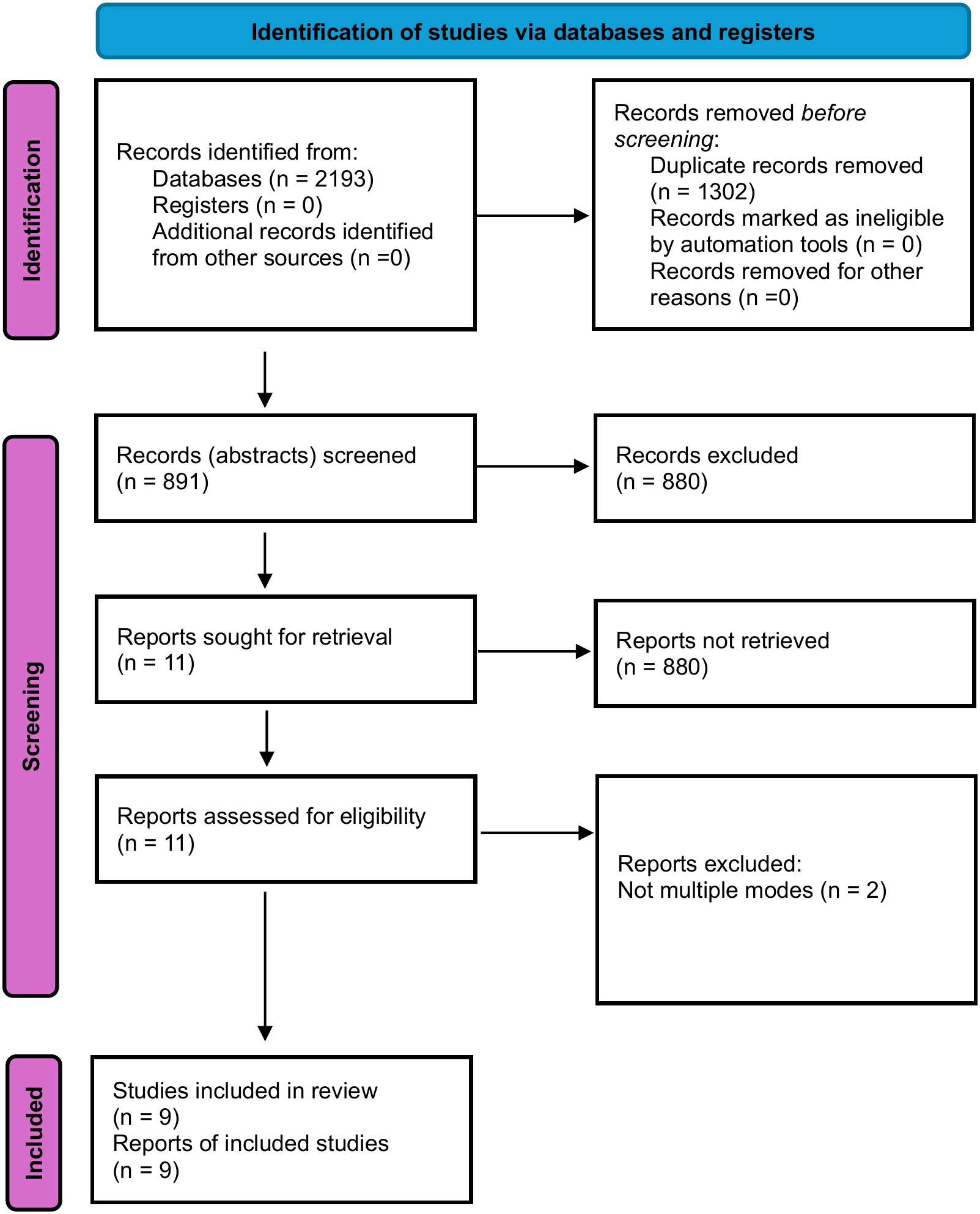
PRISMA Flow Diagram (Page et al., 2021)

### Joanna Briggs Institute (JBI) Risk of Bias Tool for Quasi-Experimental Studies

The included quasi-experimental (Q-E) studies were critically appraised for risk of bias (internal validity) by converting the responses to scores: Yes = 1, Unclear = 2, No = 3, and Not Applicable = 0. Questions with multiple responses were averaged and the range of scores is 0-27 with lower scores indicating better quality studies. A quality judgement was made according to high-quality = 0-6; moderate quality = 7-13; low quality = 14-20; and insufficient quality = 21-27. It is important to note that these judgments pertain to the comparative quality of these Q-E studies and not in relation to the overall evidence hierarchy in which Q-E studies are considered inferior to randomised controlled trials (RCTs). The quality assessment is presented in Table 3.

**Table 3.** JBI Critical Appraisal Findings.

| **Study** | **Blalock et al. (2025)** | **Carr et al. (2022)** | **Levinson et al. (2021)** | **Ortiz et al. (2023)** | **Penwell et al. (2023)** | **Raykos et al. (2021)** | **Robinson et al. (2024)** | **Steiger et al. (2021)** | **Thaler et al. (2024)** |
|---|---|---|---|---|---|---|---|---|---|
| **Total Score**<br>**(0-27)** | 11 | 14 | 13.8 | 13.6 | 12.4 | 13 | NA | 12 | 14 |
| **Risk of Bias** | Moderate | High | Moderate | Moderate | Moderate | Moderate | NA | Moderate | High |
| **QUALITY JUDGEMNET** | **MODERATE** | **LOW** | **MODERATE** | **MODERATE** | **MODERATE** | **MODERATE** | **NA** | **MODERATE** | **LOW** |

Not Applicable (NA)

### Critical Appraisal Summary

The included Q-E studies were moderate in methodological quality with only two (Carr et al., 2022; Thaler et al., 2024) being rated as low quality which is encouraging. These Q-E studies are good quality Q-E studies, but they are placed below well-designed RCTs in the evidence hierarchy (Nelson, 2025, p. 42). In the absence of RCTs, however, these studies are the best available evidence base and allow reasonable confidence in the reliability and validity of the review findings.

When interpreting the outcome of quality assessments, it is important to consider whether poor reporting was likely to influence the scores. Bias in the studies, could exaggerate or underestimate the true effect size.

In all studies: 1) temporal precedence was clear - that is the cause (interventions) came before the effect (outcomes measured); 2) selection bias and allocation bias were mitigated by using a comparison group; 3) confounding factors were reduced by ensuring participants in the comparison group were similar (i.e., from the same population-clinic); and 4) performance bias was diminished by providing similar treatment to participants in the comparison group except for the mode of treatment delivery.

The main area of identified risk was the potential for detection bias; a serious concern in all the studies, as only one set of pre-post outcome measurements were taken which makes it difficult to determine whether any changes observed are due to the intervention rather than other factors. Other forms of detection bias were attenuated by using self-report measures which are considered reliable despite respondent/recall bias issues, and both groups were assessed in the same way using patient reported outcome measures (PROMs). However, all studies which measured BMI were downrated as BMI was not measured in the same way in the comparison group; conducted onsite (in-person programmes) or by self-report (online programmes) and self-reported weighing can be inaccurate, for example, due to using home scales or from participants being deceptive about their weight.

### Study and Participants’ Characteristics

In the included studies, the mean age ranged from 19-31 years, participants were predominantly White, female, and all studies included variant EDs except Carr et al. (2022) and Robinson et al. (2024) which only included anorexia nervosa (AN) subtypes. Five of the eight studies were large studies with more than 100 participants (Blalock et al., 2025; Penwell et al., 2023; Raykos et al., 2021; Steiger et al., 2021; Thaler et al., 2024), two were medium-size studies (Levinson et al., 2021; Ortiz et al., 2023), and one can be considered as small (Carr et al., 2022). All studies were conducted in western countries (UK, Australia, Canada and the US), with the majority being US studies.

All the studies were quasi-experimental (Q-E) in design, and each compared an in-person cohort to an online cohort with only Robinson et al. (2024) including a mixed (blended/hybrid) comparison group. For one study, Blalock et al. (2025) the in-person cohort was recruited during the period when COVID-19 pandemic restrictions were lifted and compared to an online cohort treated during lockdown restrictions, at the same clinic. Due to the time overlap this study mitigates historical artefacts from disparate time periods impacting outcomes differently. The other studies compared a historical pre-pandemic in-person daycare/intensive outpatient programme cohort or outpatient cohort to a pandemic online daycare/intensive outpatient programme cohort or outpatient cohort treated at the same clinic.

These studies represent “a natural experiment” due to the forced switch to online care which allowed the evaluation of the impact of online care on all participants rather than just those who would agree to randomisation in a randomised controlled trial (RCT) or who would opt for online care, thus likely increases the generalisability of the findings. However, as participants were not randomised to the interventions, cohort effects (e.g. influence of the pandemic) and participant characteristics (e.g. treatment preference) may have affected treatment response.

Community care in higher level of care (HLOC) is based on a multidisciplinary team approach while all first-line evidence-based approaches such as cognitive behaviour therapy (CBT) and family-based treatment (FBT) tend to be outpatient care (Levinson et al., 2021). In the included studies in-person and online treatments were comparable in format although online versions tended to provide fewer hours of care per day per week. Some components of ED treatment can be difficult to deliver online such as family meals during family therapy, mirror exposure in CBT-ED, and in-session weighing (Raykos et al., 2021). In online programmes weight monitoring was typically via self-report or from a support person/general practitioner although Thaler et al. (2024) reported in-clinic weighing was done, if possible, but primarily it was virtual.

Although Penwell et al. (2023) conducted a survey at six months post-discharge it was not reported due to limited sample size. None of the studies reported follow-up on outcomes; a limitation that needs to be addressed in future research to evaluate whether treatment effects are maintained in the long-term.

Four studies did not report comorbidities (Carr et al., 2022; Ortiz et al., 2023; Steiger et al., 2021; Thaler et al., 2024), while two studies omitted information on ethnicities (Steiger et al., 2021; Thaler et al., 2024). Four studies reported a mix of ethnicities in both treatment arms (Blalock et al., 2025; Carr et al., 2022; Levinson et al., 2021; Penwell et al., 2023). Ortiz et al. (2023) had an ethnic mix in the online cohort compared to an all-White in-person cohort and Raykos et al. (2021) only reported ethnicity for the online cohort as predominantly Anglo European Australian (70%).

Data on comorbidities are required to elucidate any confounders and reporting on minority groups is critical for improving treatment for underserved populations. A range of factors such as access to better healthcare, food, and reduced stress, can result in consistently better health in high versus low socioeconomic status participants, but no study specified the socioeconomic status of participants. This is of particular concern as such data are needed to identify marginalised populations and to strive to deliver equitable, inclusive and evidence-based treatments.

Study and participants’ characteristics are presented in Table 4.

**Table 4.**
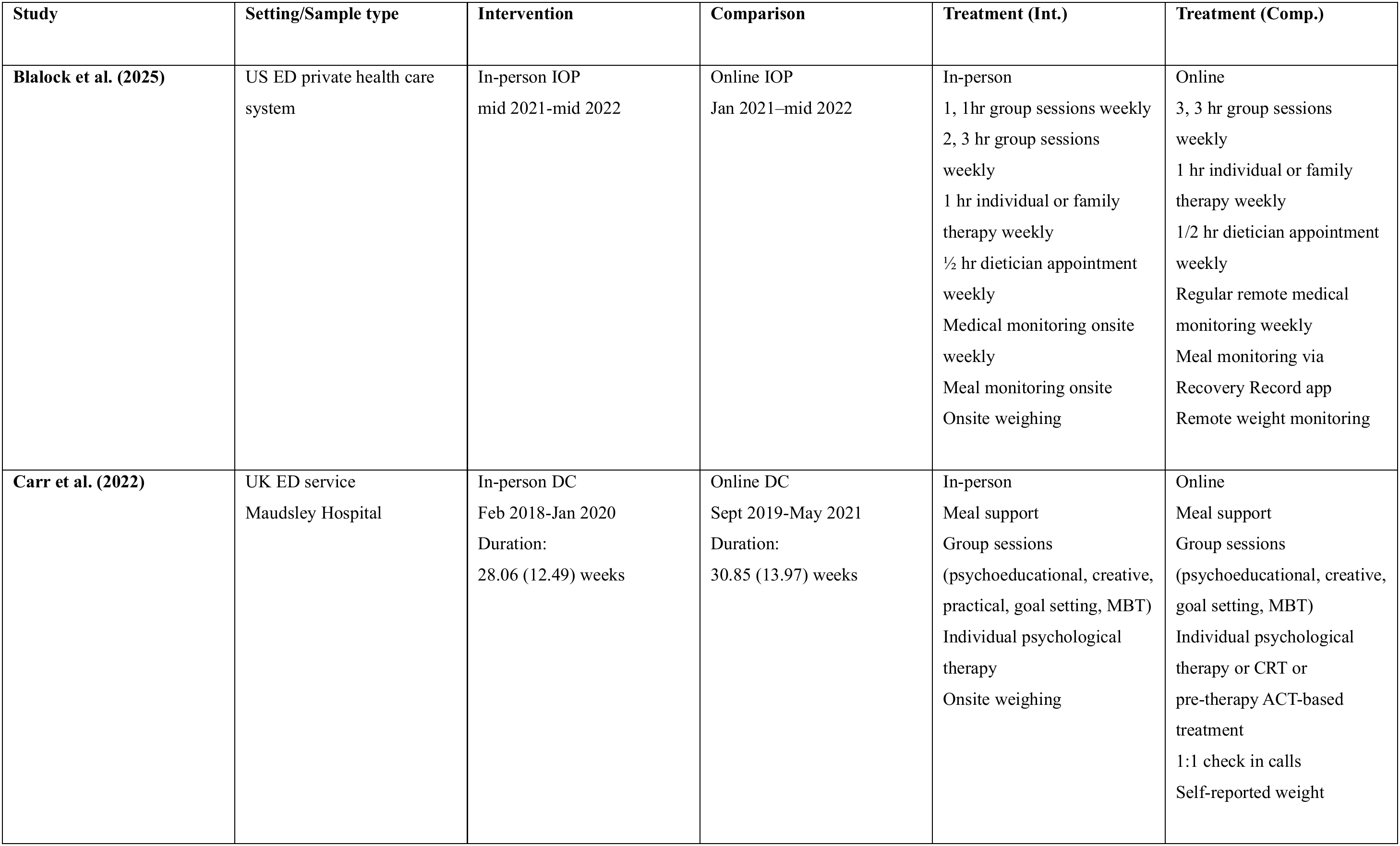
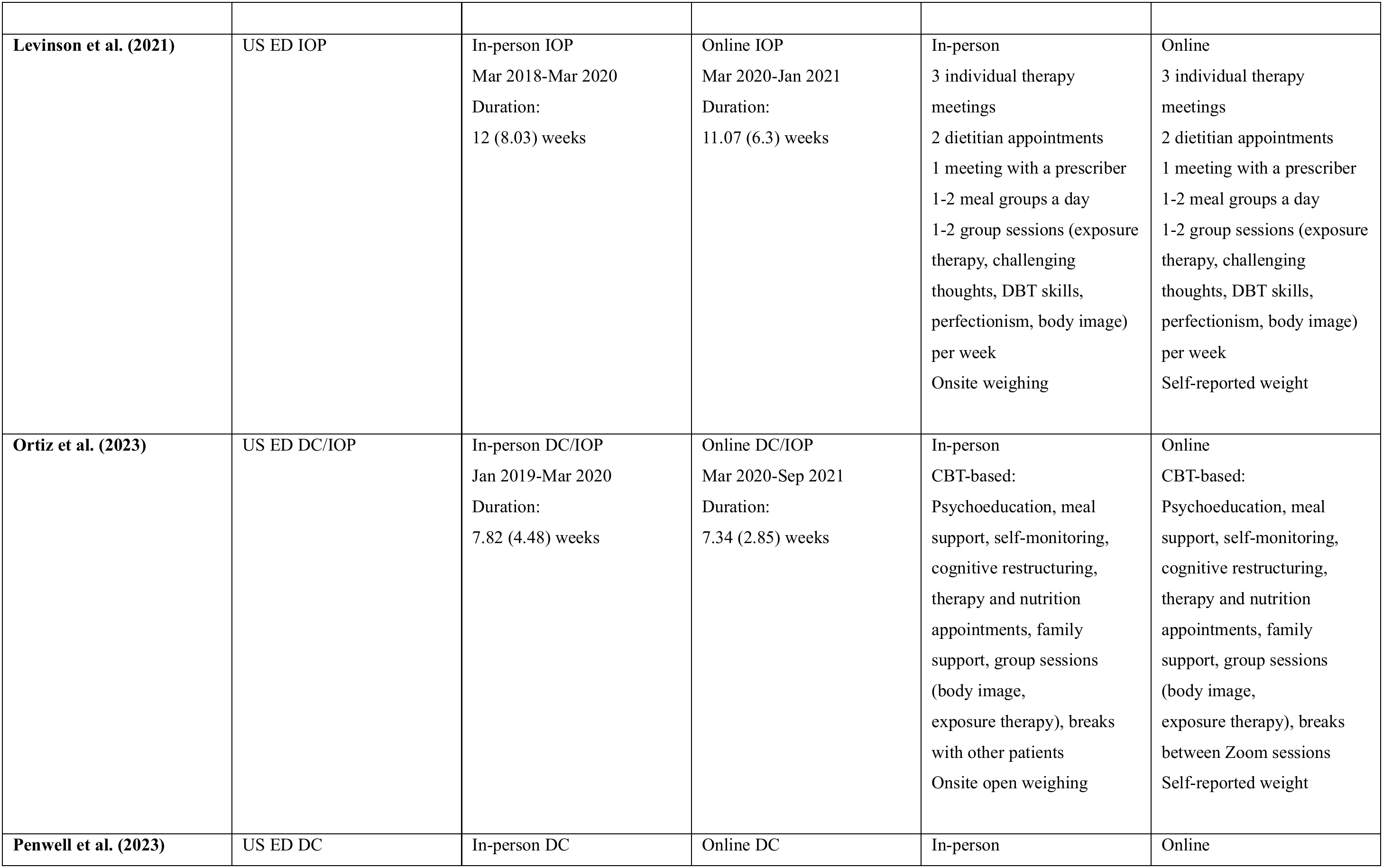
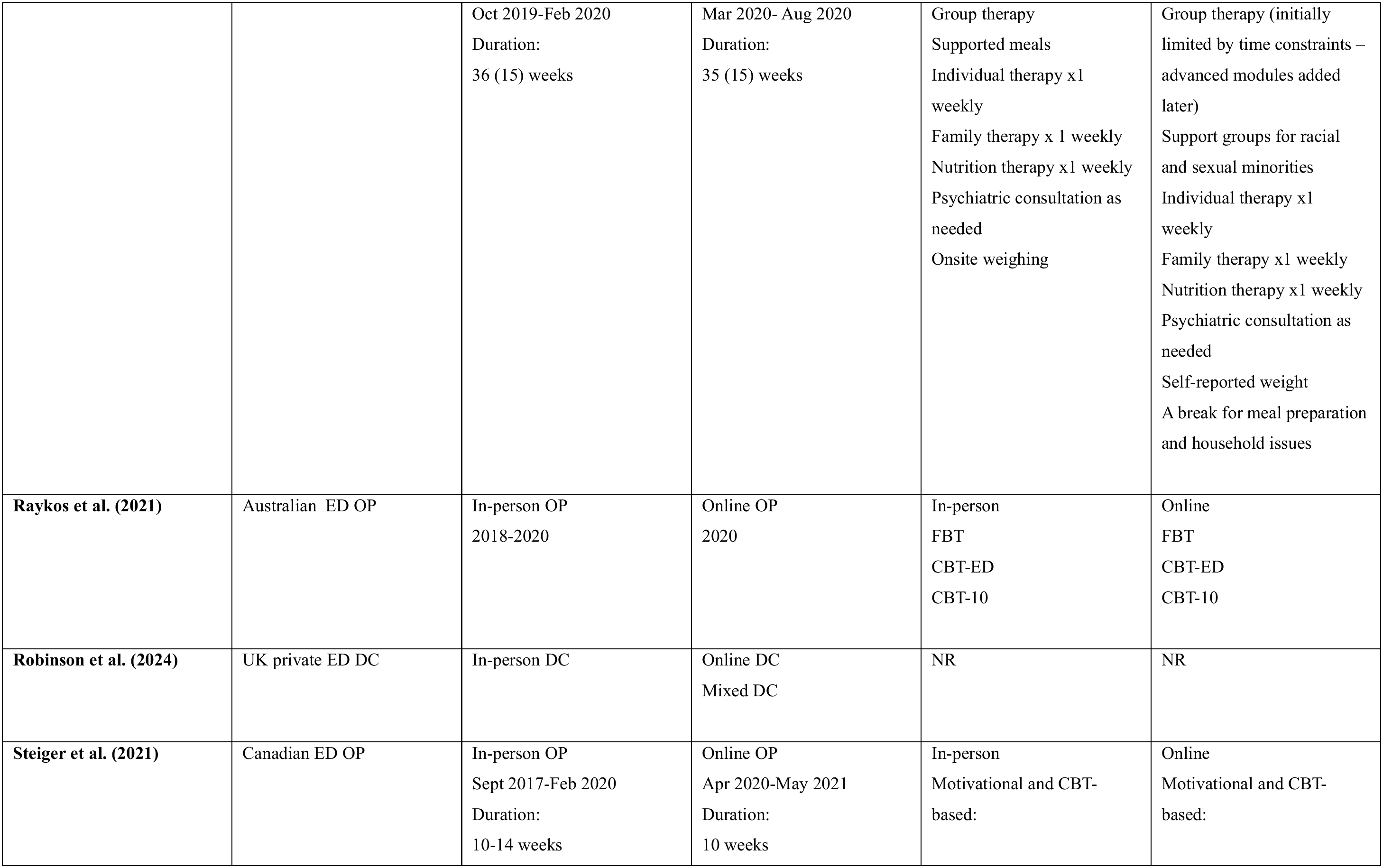
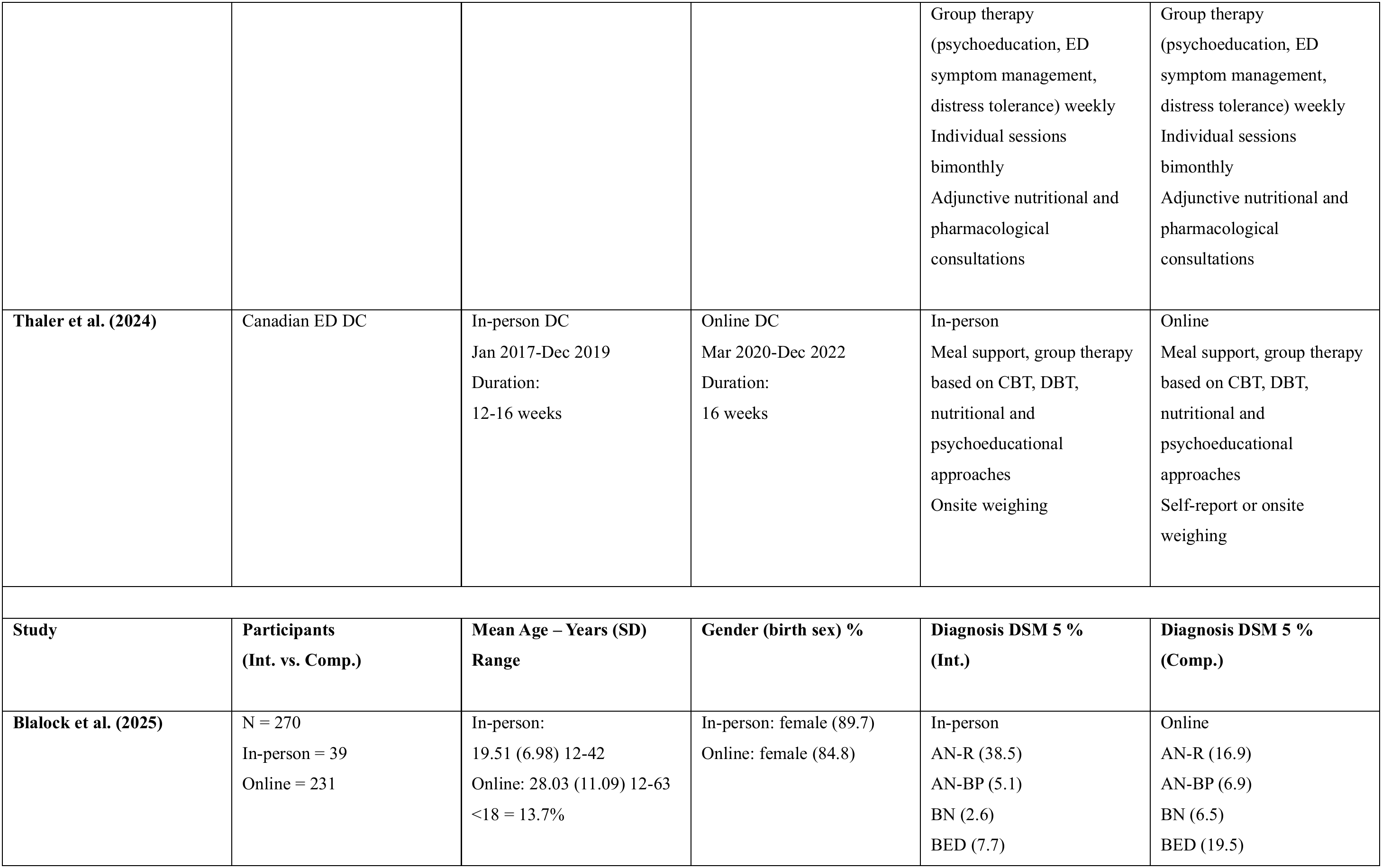
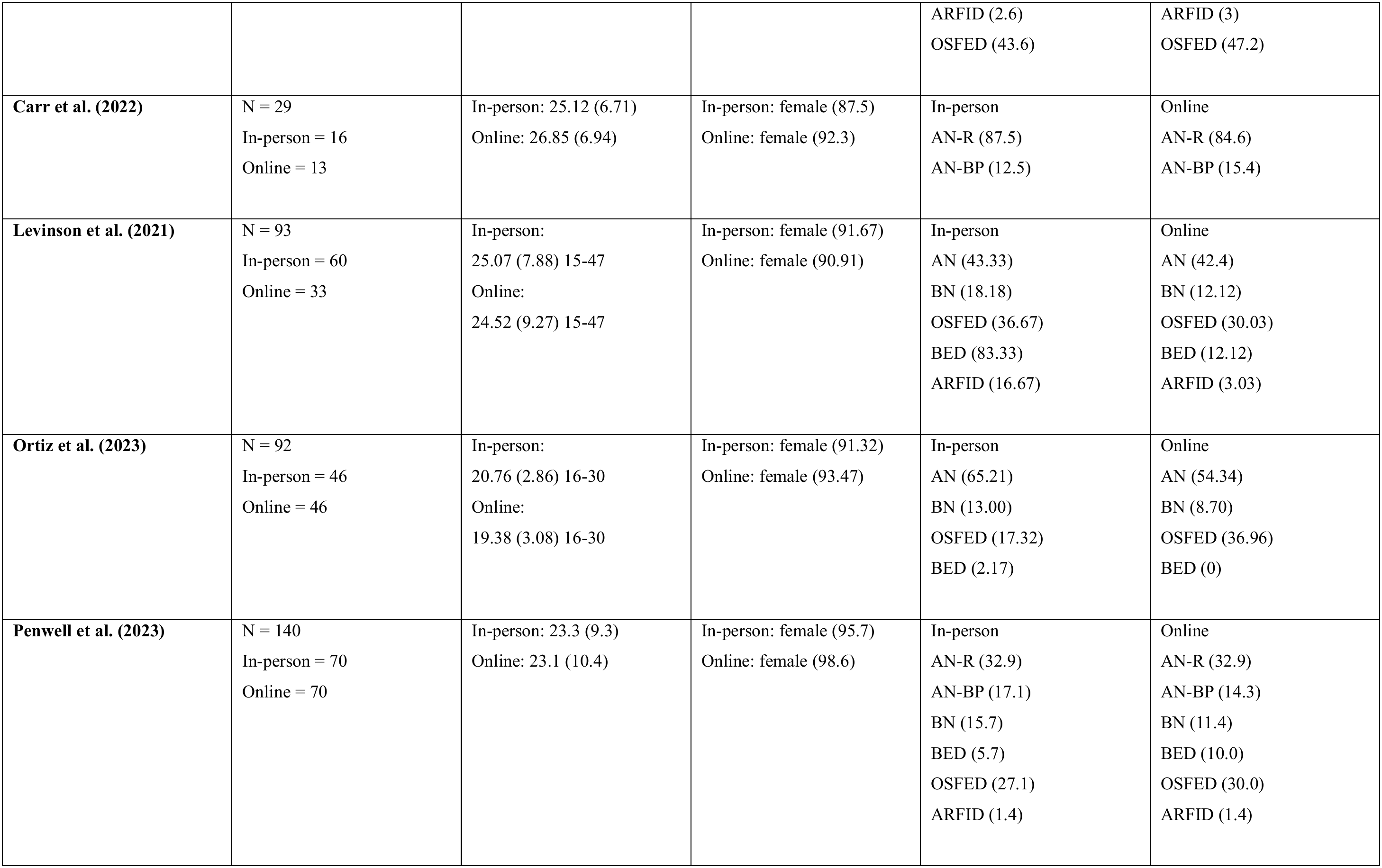
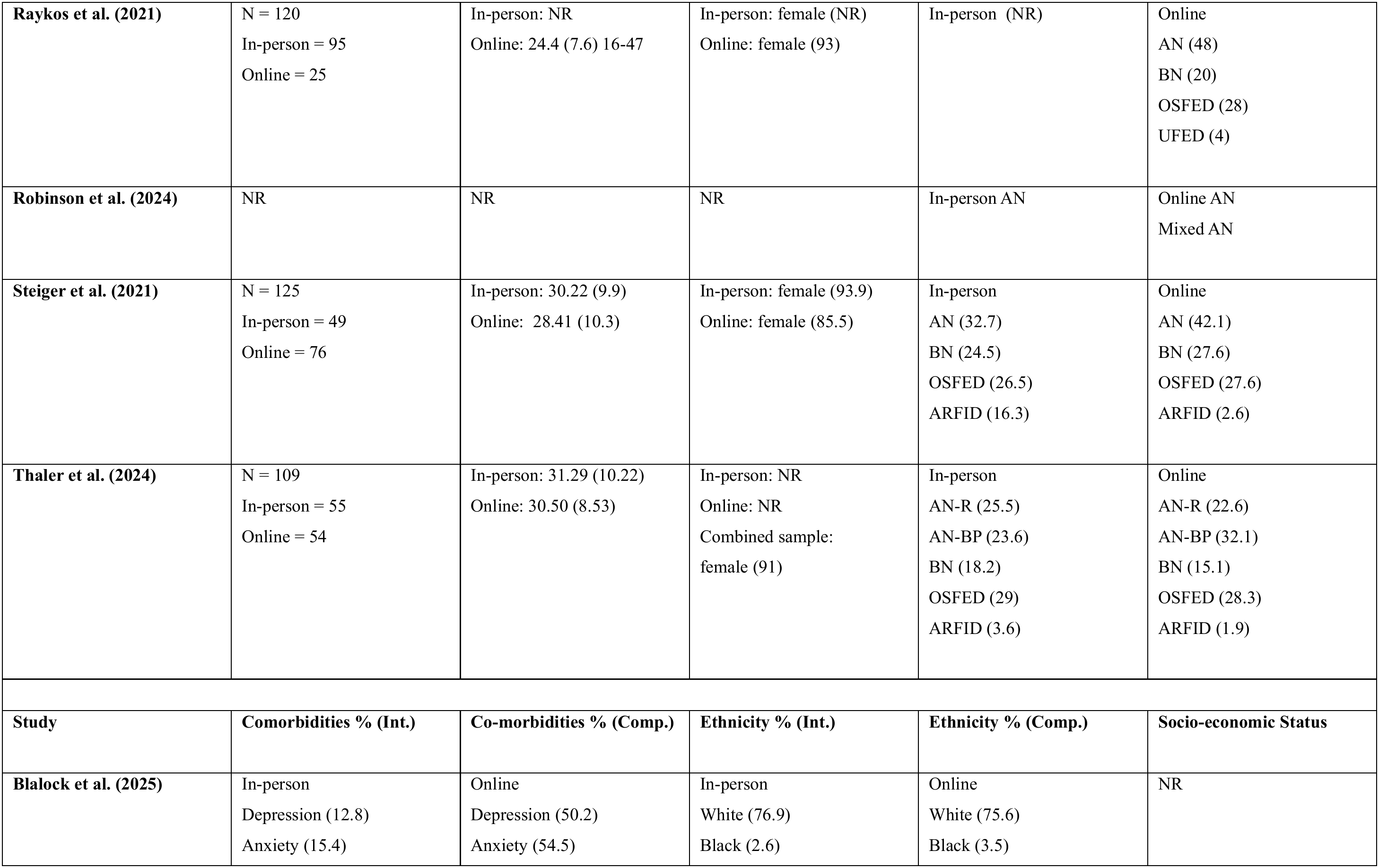
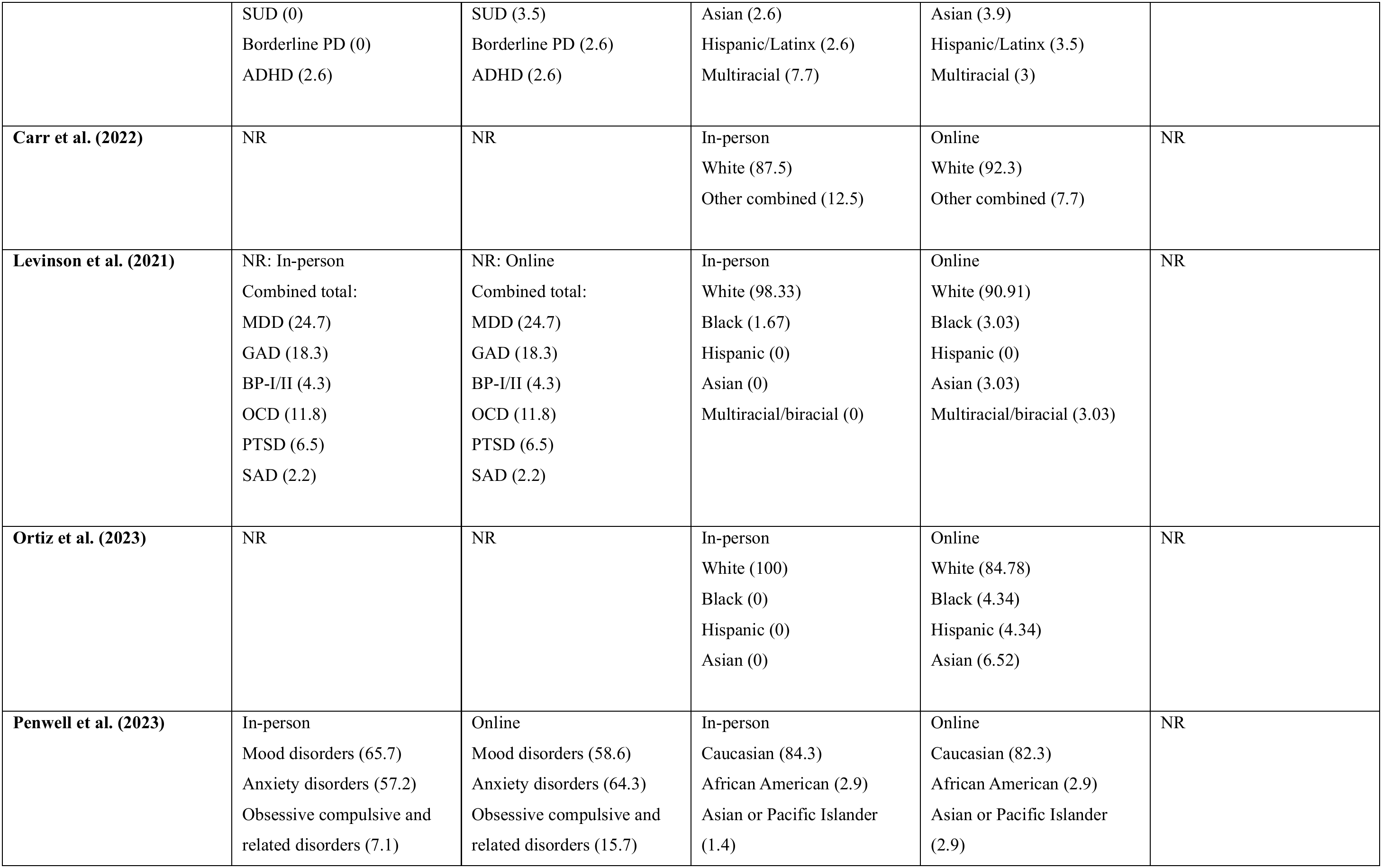
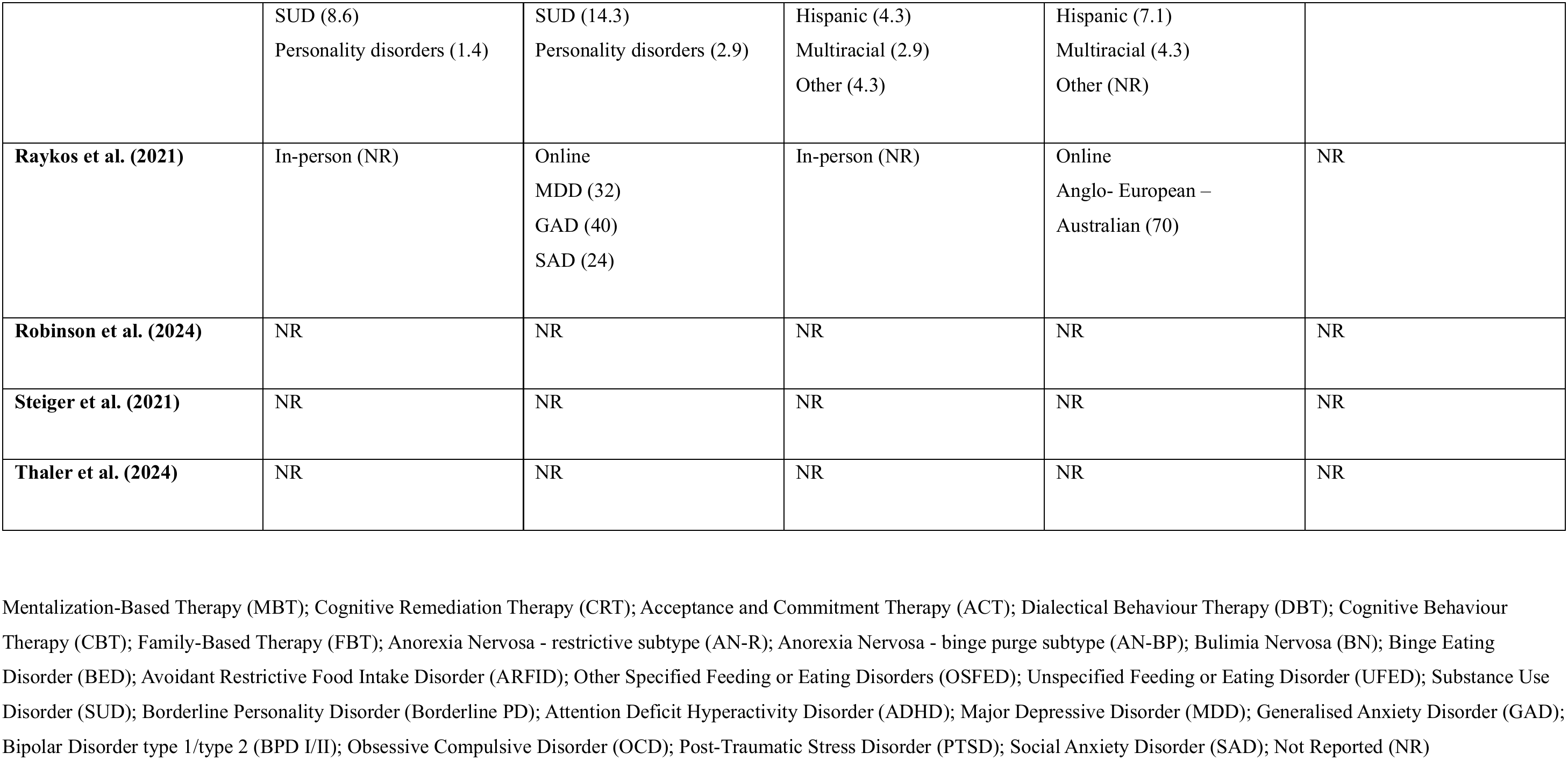
Study and Participants’ Characteristics.

| Study | Setting/Sample type | Intervention | Comparison | Treatment (Int.) | Treatment (Comp.) |
|---|---|---|---|---|---|
| **Blalock et al. (2025)** | US ED private health care system | In-person IOP<br>mid 2021-mid 2022 | Online IOP<br>Jan 2021–mid 2022 | In-person<br>1, 1hr group sessions weekly<br>2, 3 hr group sessions weekly<br>1 hr individual or family therapy weekly<br>½ hr dietician appointment weekly<br>Medical monitoring onsite weekly<br>Meal monitoring onsite<br>Onsite weighing | Online<br>3, 3 hr group sessions weekly<br>1 hr individual or family therapy weekly<br>1/2 hr dietician appointment weekly<br>Regular remote medical monitoring weekly<br>Meal monitoring via Recovery Record app<br>Remote weight monitoring |
| **Carr et al. (2022)** | UK ED service<br>Maudsley Hospital | In-person DC<br>Feb 2018-Jan 2020<br>Duration:<br>28.06 (12.49) weeks | Online DC<br>Sept 2019-May 2021<br>Duration:<br>30.85 (13.97) weeks | In-person<br>Meal support<br>Group sessions (psychoeducational, creative, practical, goal setting, MBT)<br>Individual psychological therapy<br>Onsite weighing | Online<br>Meal support<br>Group sessions (psychoeducational, creative, goal setting, MBT)<br>Individual psychological therapy or CRT or pre-therapy ACT-based treatment<br>1:1 check in calls<br>Self-reported weight |
| **Levinson et al. (2021)** | US ED IOP | In-person IOP<br>Mar 2018-Mar 2020<br>Duration:<br>12 (8.03) weeks | Online IOP<br>Mar 2020-Jan 2021<br>Duration:<br>11.07 (6.3) weeks | In-person<br>3 individual therapy meetings<br>2 dietitian appointments<br>1 meeting with a prescriber<br>1-2 meal groups a day<br>1-2 group sessions (exposure therapy, challenging thoughts, DBT skills, perfectionism, body image) per week<br>Onsite weighing | Online<br>3 individual therapy meetings<br>2 dietitian appointments<br>1 meeting with a prescriber<br>1-2 meal groups a day<br>1-2 group sessions (exposure therapy, challenging thoughts, DBT skills, perfectionism, body image) per week<br>Self-reported weight |
| **Ortiz et al. (2023)** | US ED DC/IOP | In-person DC/IOP<br>Jan 2019-Mar 2020<br>Duration:<br>7.82 (4.48) weeks | Online DC/IOP<br>Mar 2020-Sep 2021<br>Duration:<br>7.34 (2.85) weeks | In-person<br>CBT-based:<br>Psychoeducation, meal support, self-monitoring, cognitive restructuring, therapy and nutrition appointments, family support, group sessions (body image,<br>exposure therapy), breaks with other patients<br>Onsite open weighing | Online<br>CBT-based:<br>Psychoeducation, meal support, self-monitoring, cognitive restructuring, therapy and nutrition appointments, family support, group sessions (body image,<br>exposure therapy), breaks between Zoom sessions<br>Self-reported weight |
| **Penwell et al. (2023)** | US ED DC | In-person DC<br>Oct 2019-Feb 2020<br>Duration:<br>36 (15) weeks | Online DC<br>Mar 2020- Aug 2020<br>Duration:<br>35 (15) weeks | In-person<br>Group therapy<br>Supported meals<br>Individual therapy x1 weekly<br>Family therapy x 1 weekly<br>Nutrition therapy x1 weekly<br>Psychiatric consultation as needed<br>Onsite weighing | Online<br>Group therapy (initially limited by time constraints – advanced modules added later)<br>Support groups for racial and sexual minorities<br>Individual therapy x1 weekly<br>Family therapy x1 weekly<br>Nutrition therapy x1 weekly<br>Psychiatric consultation as needed<br>Self-reported weight<br>A break for meal preparation and household issues |
| **Raykos et al. (2021)** | Australian ED OP | In-person OP<br>2018-2020 | Online OP<br>2020 | In-person<br>FBT<br>CBT-ED<br>CBT-10 | Online<br>FBT<br>CBT-ED<br>CBT-10 |
| **Robinson et al. (2024)** | UK private ED DC | In-person DC | Online DC<br>Mixed DC | NR | NR |
| **Steiger et al. (2021)** | Canadian ED OP | In-person OP<br>Sept 2017-Feb 2020<br>Duration:<br>10-14 weeks | Online OP<br>Apr 2020-May 2021<br>Duration:<br>10 weeks | In-person<br>Motivational and CBT-based:<br>Group therapy (psychoeducation, ED symptom management, distress tolerance) weekly<br>Individual sessions bimonthly<br>Adjunctive nutritional and pharmacological consultations | Online<br>Motivational and CBT-based:<br>Group therapy (psychoeducation, ED symptom management, distress tolerance) weekly<br>Individual sessions bimonthly<br>Adjunctive nutritional and pharmacological consultations |
| **Thaler et al. (2024)** | Canadian ED DC | In-person DC<br>Jan 2017-Dec 2019<br>Duration:<br>12-16 weeks | Online DC<br>Mar 2020-Dec 2022<br>Duration:<br>16 weeks | In-person<br>Meal support, group therapy based on CBT, DBT, nutritional and psychoeducational approaches<br>Onsite weighing | Online<br>Meal support, group therapy based on CBT, DBT, nutritional and psychoeducational approaches<br>Self-report or onsite weighing |
| **Study** | **Participants**<br>**(Int. vs. Comp.)** | **Mean Age – Years (SD)**<br>**Range** | **Gender (birth sex) %** | **Diagnosis DSM 5 %**<br>**(Int.)** | **Diagnosis DSM 5 %**<br>**(Comp.)** |
| **Blalock et al. (2025)** | N = 270<br>In-person = 39<br>Online = 231 | In-person:<br>19.51 (6.98) 12-42<br>Online: 28.03 (11.09) 12-63<br><18 = 13.7% | In-person: female (89.7)<br>Online: female (84.8) | In-person<br>AN-R (38.5)<br>AN-BP (5.1)<br>BN (2.6)<br>BED (7.7)<br>ARFID (2.6)<br>OSFED (43.6) | Online<br>AN-R (16.9)<br>AN-BP (6.9)<br>BN (6.5)<br>BED (19.5)<br>ARFID (3)<br>OSFED (47.2) |
| **Carr et al. (2022)** | N = 29<br>In-person = 16<br>Online = 13 | In-person: 25.12 (6.71)<br>Online: 26.85 (6.94) | In-person: female (87.5)<br>Online: female (92.3) | In-person<br>AN-R (87.5)<br>AN-BP (12.5) | Online<br>AN-R (84.6)<br>AN-BP (15.4) |
| **Levinson et al. (2021)** | N = 93<br>In-person = 60<br>Online = 33 | In-person:<br>25.07 (7.88) 15-47<br>Online:<br>24.52 (9.27) 15-47 | In-person: female (91.67)<br>Online: female (90.91) | In-person<br>AN (43.33)<br>BN (18.18)<br>OSFED (36.67)<br>BED (83.33)<br>ARFID (16.67) | Online<br>AN (42.4)<br>BN (12.12)<br>OSFED (30.03)<br>BED (12.12)<br>ARFID (3.03) |
| **Ortiz et al. (2023)** | N = 92<br>In-person = 46<br>Online = 46 | In-person:<br>20.76 (2.86) 16-30<br>Online:<br>19.38 (3.08) 16-30 | In-person: female (91.32)<br>Online: female (93.47) | In-person<br>AN (65.21)<br>BN (13.00)<br>OSFED (17.32)<br>BED (2.17) | Online<br>AN (54.34)<br>BN (8.70)<br>OSFED (36.96)<br>BED (0) |
| **Penwell et al. (2023)** | N = 140<br>In-person = 70<br>Online = 70 | In-person: 23.3 (9.3)<br>Online: 23.1 (10.4) | In-person: female (95.7)<br>Online: female (98.6) | In-person<br>AN-R (32.9)<br>AN-BP (17.1)<br>BN (15.7)<br>BED (5.7)<br>OSFED (27.1)<br>ARFID (1.4) | Online<br>AN-R (32.9)<br>AN-BP (14.3)<br>BN (11.4)<br>BED (10.0)<br>OSFED (30.0)<br>ARFID (1.4) |
| **Raykos et al. (2021)** | N = 120<br>In-person = 95<br>Online = 25 | In-person: NR<br>Online: 24.4 (7.6) 16-47 | In-person: female (NR)<br>Online: female (93) | In-person (NR) | Online<br>AN (48)<br>BN (20)<br>OSFED (28)<br>UFED (4) |
| **Robinson et al. (2024)** | NR | NR | NR | In-person AN | Online AN<br>Mixed AN |
| **Steiger et al. (2021)** | N = 125<br>In-person = 49<br>Online = 76 | In-person: 30.22 (9.9)<br>Online: 28.41 (10.3) | In-person: female (93.9)<br>Online: female (85.5) | In-person<br>AN (32.7)<br>BN (24.5)<br>OSFED (26.5)<br>ARFID (16.3) | Online<br>AN (42.1)<br>BN (27.6)<br>OSFED (27.6)<br>ARFID (2.6) |
| **Thaler et al. (2024)** | N = 109<br>In-person = 55<br>Online = 54 | In-person: 31.29 (10.22)<br>Online: 30.50 (8.53) | In-person: NR<br>Online: NR<br>Combined sample: female (91) | In-person<br>AN-R (25.5)<br>AN-BP (23.6)<br>BN (18.2)<br>OSFED (29)<br>ARFID (3.6) | Online<br>AN-R (22.6)<br>AN-BP (32.1)<br>BN (15.1)<br>OSFED (28.3)<br>ARFID (1.9) |
| **Study** | **Comorbidities % (Int.)** | **Co-morbidities % (Comp.)** | **Ethnicity % (Int.)** | **Ethnicity % (Comp.)** | **Socio-economic Status** |
| **Blalock et al. (2025)** | In-person<br>Depression (12.8)<br>Anxiety (15.4)<br>SUD (0)<br>Borderline PD (0)<br>ADHD (2.6) | Online<br>Depression (50.2)<br>Anxiety (54.5)<br>SUD (3.5)<br>Borderline PD (2.6)<br>ADHD (2.6) | In-person<br>White (76.9)<br>Black (2.6)<br>Asian (2.6)<br>Hispanic/Latinx (2.6)<br>Multiracial (7.7) | Online<br>White (75.6)<br>Black (3.5)<br>Asian (3.9)<br>Hispanic/Latinx (3.5)<br>Multiracial (3) | NR |
| **Carr et al. (2022)** | NR | NR | In-person<br>White (87.5)<br>Other combined (12.5) | Online<br>White (92.3)<br>Other combined (7.7) | NR |
| **Levinson et al. (2021)** | NR: In-person<br>Combined total:<br>MDD (24.7)<br>GAD (18.3)<br>BP-I/II (4.3)<br>OCD (11.8)<br>PTSD (6.5)<br>SAD (2.2) | NR: Online<br>Combined total:<br>MDD (24.7)<br>GAD (18.3)<br>BP-I/II (4.3)<br>OCD (11.8)<br>PTSD (6.5)<br>SAD (2.2) | In-person<br>White (98.33)<br>Black (1.67)<br>Hispanic (0)<br>Asian (0)<br>Multiracial/biracial (0) | Online<br>White (90.91)<br>Black (3.03)<br>Hispanic (0)<br>Asian (3.03)<br>Multiracial/biracial (3.03) | NR |
| **Ortiz et al. (2023)** | NR | NR | In-person<br>White (100)<br>Black (0)<br>Hispanic (0)<br>Asian (0) | Online<br>White (84.78)<br>Black (4.34)<br>Hispanic (4.34)<br>Asian (6.52) | NR |
| **Penwell et al. (2023)** | In-person<br>Mood disorders (65.7)<br>Anxiety disorders (57.2)<br>Obsessive compulsive and related disorders (7.1)<br>SUD (8.6)<br>Personality disorders (1.4) | Online<br>Mood disorders (58.6)<br>Anxiety disorders (64.3)<br>Obsessive compulsive and related disorders (15.7)<br>SUD (14.3)<br>Personality disorders (2.9) | In-person<br>Caucasian (84.3)<br>African American (2.9)<br>Asian or Pacific Islander (1.4)<br>Hispanic (4.3)<br>Multiracial (2.9)<br>Other (4.3) | Online<br>Caucasian (82.3)<br>African American (2.9)<br>Asian or Pacific Islander (2.9)<br>Hispanic (7.1)<br>Multiracial (4.3)<br>Other (NR) | NR |
| **Raykos et al. (2021)** | In-person (NR) | Online<br>MDD (32)<br>GAD (40)<br>SAD (24) | In-person (NR) | Online<br>Anglo- European – Australian (70) | NR |
| **Robinson et al. (2024)** | NR | NR | NR | NR | NR |
| **Steiger et al. (2021)** | NR | NR | NR | NR | NR |
| **Thaler et al. (2024)** | NR | NR | NR | NR | NR |

Mentalization-Based Therapy (MBT); Cognitive Remediation Therapy (CRT); Acceptance and Commitment Therapy (ACT); Dialectical Behaviour Therapy (DBT); Cognitive Behaviour Therapy (CBT); Family-Based Therapy (FBT); Anorexia Nervosa - restrictive subtype (AN-R); Anorexia Nervosa - binge purge subtype (AN-BP); Bulimia Nervosa (BN); Binge Eating Disorder (BED); Avoidant Restrictive Food Intake Disorder (ARFID); Other Specified Feeding or Eating Disorders (OSFED); Unspecified Feeding or Eating Disorder (UFED); Substance Use Disorder (SUD); Borderline Personality Disorder (Borderline PD); Attention Deficit Hyperactivity Disorder (ADHD); Major Depressive Disorder (MDD); Generalised Anxiety Disorder (GAD); Bipolar Disorder type 1/type 2 (BPD I/II); Obsessive Compulsive Disorder (OCD); Post-Traumatic Stress Disorder (PTSD); Social Anxiety Disorder (SAD); Not Reported (NR)

### Meta-Analyses

Statistically significant summary effect sizes (p ≤ 0.05), extent of heterogeneity (I^2^), and GRADE ratings for the included studies are presented in Table 5.

**Table 5.**
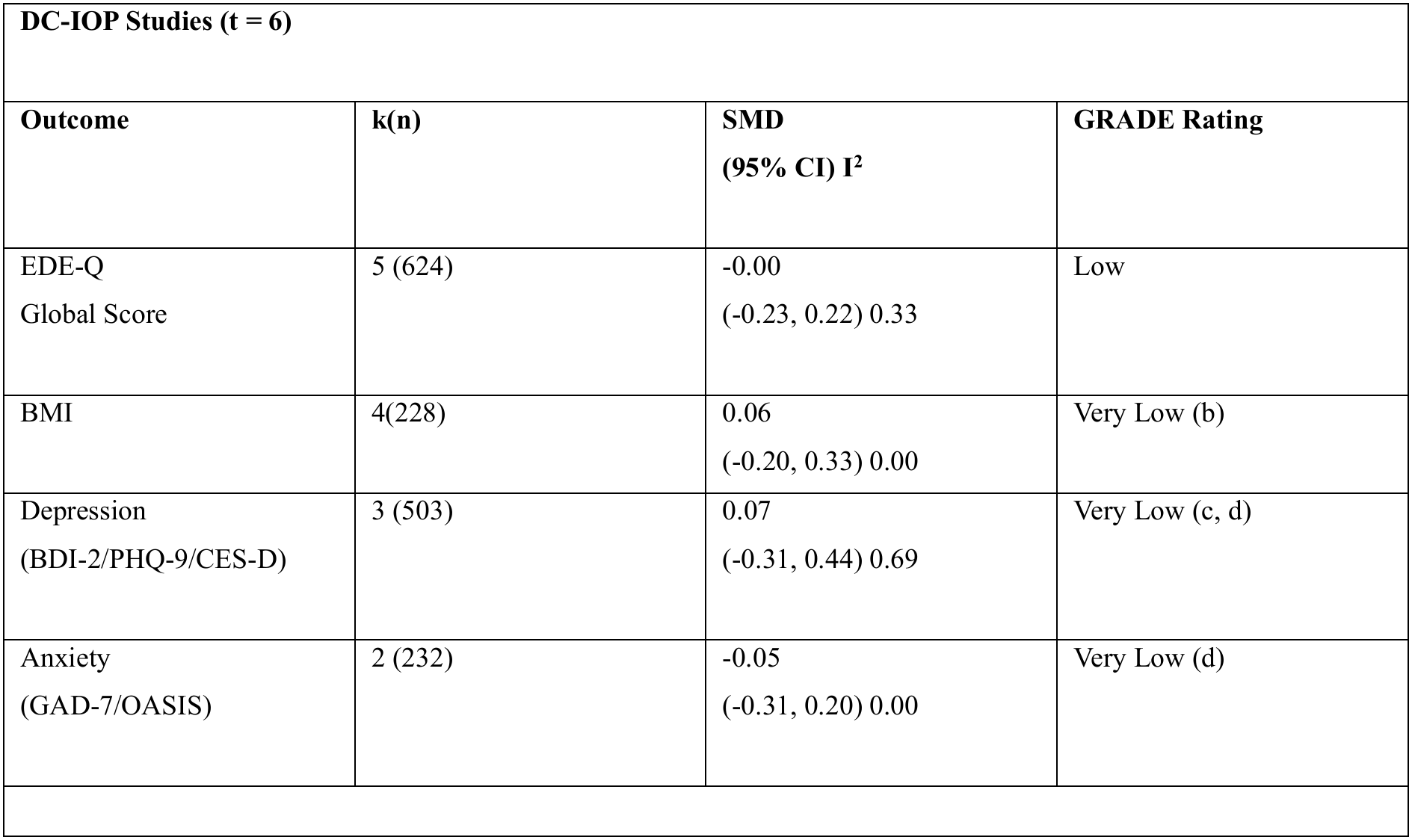
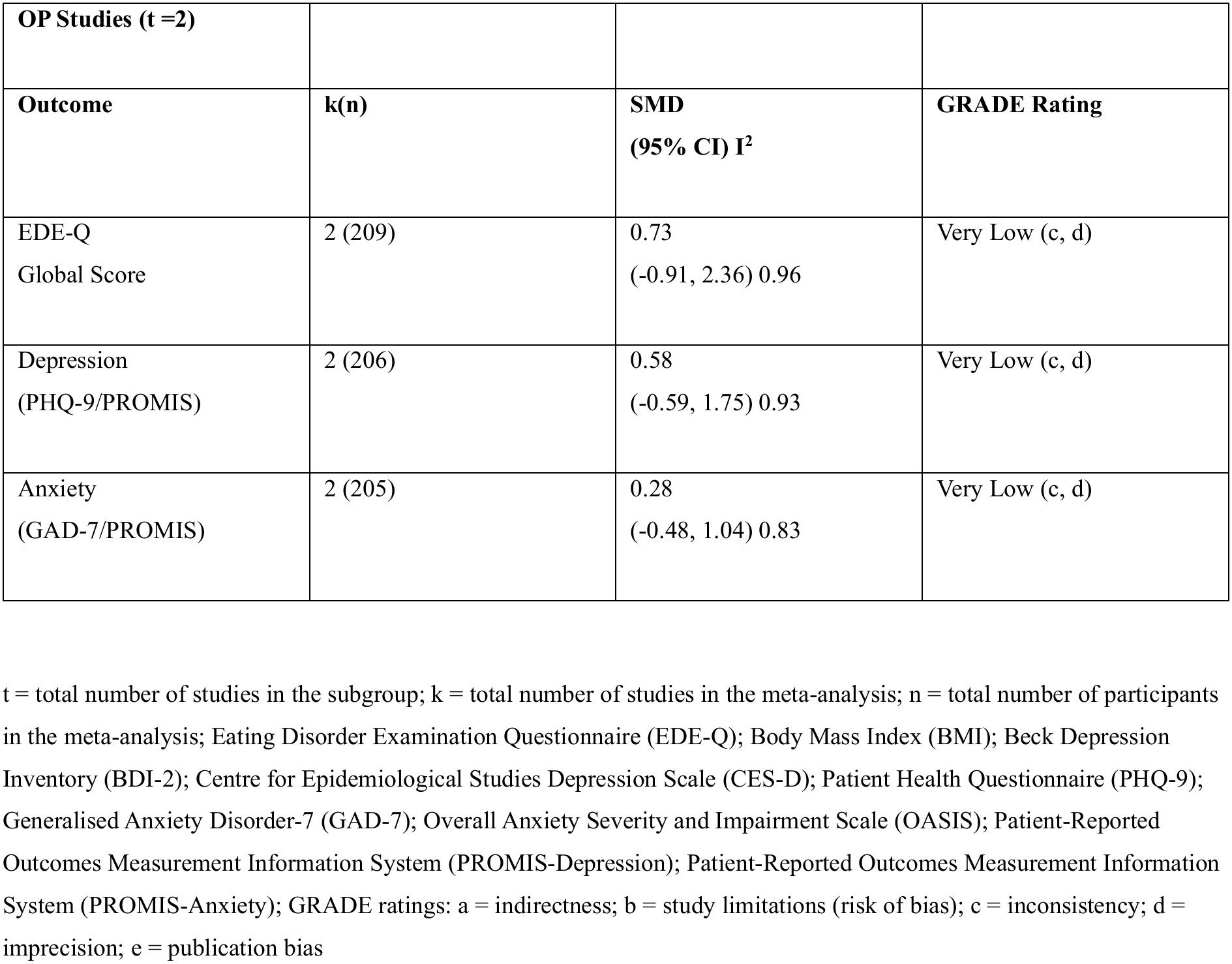
Meta-Analyses (in-person vs. online comparisons)

| **DC-IOP Studies (t = 6)** |  |  |  |
| **Outcome** | **k(n)** | **SMD**<br>**(95% CI) I<sup>2</sup>** | **GRADE Rating** |
|---|---|---|---|
| EDE-Q<br>Global Score | 5 (624) | -0.00<br>(-0.23, 0.22) 0.33 | Low |
| BMI | 4(228) | 0.06<br>(-0.20, 0.33) 0.00 | Very Low (b) |
| Depression<br>(BDI-2/PHQ-9/CES-D) | 3 (503) | 0.07<br>(-0.31, 0.44) 0.69 | Very Low (c, d) |
| Anxiety<br>(GAD-7/OASIS) | 2 (232) | -0.05<br>(-0.31, 0.20) 0.00 | Very Low (d) |
| **OP Studies (t =2)** |  |  |  |
| **Outcome** | **k(n)** | **SMD**<br>**(95% CI) I<sup>2</sup>** | **GRADE Rating** |
| EDE-Q<br>Global Score | 2 (209) | 0.73<br>(-0.91, 2.36) 0.96 | Very Low (c, d) |
| Depression<br>(PHQ-9/PROMIS) | 2 (206) | 0.58<br>(-0.59, 1.75) 0.93 | Very Low (c, d) |
| Anxiety<br>(GAD-7/PROMIS) | 2 (205) | 0.28<br>(-0.48, 1.04) 0.83 | Very Low (c, d) |

t = total number of studies in the subgroup; k = total number of studies in the meta-analysis; n = total number of participants in the meta-analysis; Eating Disorder Examination Questionnaire (EDE-Q); Body Mass Index (BMI); Beck Depression Inventory (BDI-2); Centre for Epidemiological Studies Depression Scale (CES-D); Patient Health Questionnaire (PHQ-9); Generalised Anxiety Disorder-7 (GAD-7); Overall Anxiety Severity and Impairment Scale (OASIS); Patient-Reported Outcomes Measurement Information System (PROMIS-Depression); Patient-Reported Outcomes Measurement Information System (PROMIS-Anxiety); GRADE ratings: a = indirectness; b = study limitations (risk of bias); c = inconsistency; d = imprecision; e = publication bias

Change-from-baseline between group effect size, standardised mean difference (SMD), in this case Hedges’ (adjusted) *g* was the effect size used in the meta-analyses of the daycare/intensive outpatient programme (DC-IOP) studies. As the outpatient treatment (OP) studies did not provide the required data only post-treatment between group effect size (without change scores) could be calculated which needs extra caution in interpretation of the results as baseline comparability cannot be assumed in quasi-experimental (Q-E) studies.

A null value of zero shows equivalence for the two groups. The 95% confidence interval (CI) is the range within which the true value can be found 95% of the time. If the CI includes the null value, the effect estimate is not significant (p > 0.05). When the CI excludes the null value, the effect estimate is statistically significant (p ≤ 0.05); it is unlikely to be due to chance and the null hypothesis of no difference between the groups can be rejected. Largely (or moderately) overlapping CIs suggest that estimates are similar enough for a summary estimate to be appropriate.

As evident from Table 5 for the DC-IOP studies the effect sizes for the differences between the intervention (in-person) and comparison (online) groups fall below the non-inferiority margin set at Hedges’ *g* < 0.1 and the effect sizes are not statistically significant. Although the 95% CIs are broader than *g* = +/−0.1, the two treatment delivery modes can still arguably be considered non-inferior (comparable) in effectiveness showing similar improvements in eating disorder symptoms (EDE-Q Global score), BMI, depression and anxiety symptoms.

For the OP studies posttreatment between group effect sizes (without change scores) were calculated which do not account for baseline variability and need to be interpreted with caution. Table 5 shows that the effect sizes for the differences between the intervention (in-person) and comparison (online) groups do not fall below the non-inferiority margin set at Hedges’ *g* < 0.1 but the effect sizes are not statistically significant so the null hypothesis of equivalence between the two groups cannot be rejected based on these results. Thus, the two treatment delivery modes can be considered comparable in effectiveness showing similar improvements in eating disorder symptoms (EDE-Q Global score), depression and anxiety symptoms. However, compared to the DC-IOP studies the OP studies showed larger between group differences, though non-significant, indicating possible type 2 error.

*Sensitivity analyses* showed robustness of the pooled estimates regarding study quality and type of ED. *Heterogeneity* was more of a concern for the OP studies and could be improved by using change-from-baseline between group effect sizes and by comparing similar OP treatments with homogenous participants (e.g., matching severity and type of ED). Analyses undertaken reinforced that *publication bias* was not of great concern in this study and the search strategy was exhaustive, but searching grey literature would have increased certainty in the findings. These results are limited by the small number of included studies. The *GRADE ratings* for the outcomes were mostly very low and decrease the credibility of the results. Despite the drawbacks, the results of this systematic review and meta-analysis are likely to be the best in the field so far as the data are derived exclusively from QE studies - the best evidence base currently available.

## DISCUSSION

### Is Telemedicine a Viable Option?

#### Narrative Synthesis

This systematic review and meta-analysis aimed to compare the effectiveness of in-person vs. online vs. mixed (blended/hybrid) modes of ED community care delivery for adults globally.

The majority of the included studies were of moderate methodological quality (internal validity) despite low-very low, GRADE ratings. Certainty in the findings is attenuated by the small number of included studies, but despite several studies being underpowered all studies used reliable and valid patient reported outcome measures, thus increasing the credibility of the research.

Recruitment bias was mitigated by selecting participants in both groups from the same population (ED clinic). However, as participants were recruited during different time periods there may be confounding by temporal factors, in particular the COVID-19 pandemic context in all studies for the online cohorts; results may be different in non-pandemic cohorts and generalisability despite a mix of ethnicities in most studies is limited to a predominantly White, female, and high-income western population.

Co-morbidities were present in the samples allowing some generalisability to a broader population. But studies did not report whether participants were receiving any additional pharmacological or psychotherapeutic treatments outside of the study which could have confounded the findings.

Compliance with the programmes, a possible confounder was described by all but one study (Raykos et al., 2021) and can be considered moderate indicating that these treatments are acceptable to participants. Improving compliance could lead to better outcomes. Factors affecting compliance such as baseline ED symptom severity; type of ED diagnosis; information technology (IT) difficulties; absence of face-to-face contact; confidentiality/patient privacy and data security issues need to be evaluated. Monetary incentives were not offered in any of the studies which aligns with routine clinical care, but adding motivational interviewing components could improve adherence and ultimately the outcomes.

Adverse effects were not reported in any study. This lack of reporting may be concealing adverse events, but on the contrary, does imply that the interventions did not have any negative effects; patient safety would need clarification before dissemination. In a service evaluation of an ED DC programme for adults in Surrey (UK), Obiekezie et al. (2022) reported that online and mixed (blended/hybrid) DC, for a sample of AN patients with either subtype or unspecified AN, was relatively safe alongside weekly in-person physical health monitoring and regular health investigations to manage risks to health.

Two possible confounders that need to be considered are history of ED and past ED treatment which were not described in the included studies. A negative past experience of ED or its treatment may reduce expectations for successful treatment and adversely bias the results. Similarly, prior experience with and expectations of online/mixed treatment were not accounted for and could affect treatment impact.

Follow-up was not reported in any included study which is a critical limitation. It is suggested that a follow-up of at least six months is required to clarify whether any benefits from the treatments are sustained and at least two to three years follow-up is necessary to assess whether the programmes increase abstinence from an ED in this study population.

#### Meta-Analyses

The daycare/intensive outpatient programme (DC-IOP) studies can be considered as showing non-inferiority of in-person and online treatment programmes: EDE-Q Global score (−0.00 [95% CI: −0.23, 0.22]); anxiety (GAD-7/OASIS) (−0.05 [95% CI: −.31, 0.20]); BMI (0.06 [95% CI: −0.20, 0.33]); and depression (BDI-2/PHQ-9/CES-D) (0.07 [95% CI: −0.31, 0.44]) all fall well below the non-inferiority margin set at Hedges’ *g* < 0.1 although the 95% CIs are broader than *g* = +/−0.1. Further, these effect sizes are not statistically significant (i.e. the null hypothesis of no difference cannot be rejected).

The outpatient treatment (OP) studies did not show non-inferiority of in-person and online treatment programmes: anxiety (GAD-7/PROMIS) (0.28 [95% CI: −0.48, 1.04]); depression (PHQ-9/PROMIS) (0.58 [95% CI: −0.59, 1.75]); and EDE-Q Global score (0.73 [95% CI: - 0.91, 2.36]) all fall above the non-inferiority margin set at Hedges’ *g* < 0.1 but the effect sizes are not statistically significant (i.e. the null hypothesis of no difference cannot be rejected). For depression and ED symptoms differences exceeded the minimally important difference, MID: SMD = 0.5 (Norman et al., 2003); possibly with larger sample sizes these differences could reach statistical significance thus would be clinically important. It must be noted that most of the effect sizes in the OP studies were very imprecise as evident from extremely wide confidence intervals.

The null hypothesis of equivalence between the two groups cannot be rejected based on these meta-analyses results. Thus, the two treatment delivery modes can be considered comparable in effectiveness showing similar improvements in eating disorder symptoms (EDE-Q Global score), BMI, depression and anxiety symptoms. However, compared to the DC-IOP studies the OP studies showed larger between group differences which were non-significant indicating possible type 2 error. Moreover, the DC-IOP studies were analysed using change scores while for the OP studies posttreatment between group effect sizes (without change scores) were used which do not account for baseline variability.

For the DC-IOP studies by considering the direction of the effects it is evident that there is slightly or marginally more improvement in anxiety and BMI respectively, in the in-person group, while the online group showed slightly more improvement in depression symptoms. ED symptoms (EDE-Q Global score), however, showed similar benefits for both treatment delivery modes. Anxiety may have been elevated in the online cohort due to the context of the COVID-19 pandemic and BMI may have been inaccurate in the online group as a consequence of reduced accountability allowing for deception/falsification by patients and from using unreliable home scales for weight monitoring. Similarly, the pandemic context could have reduced depression symptoms in the online group due to families being isolated together and having more time to support family members with an ED.

Examining the direction of effects for the OP studies shows slightly more improvement in the online group than in the in-person group on all measures: eating disorder symptoms (EDE-Q Global score), depression and anxiety symptoms. An explanation for the enhanced benefits of online OP treatment may be the pandemic context resulting in more support from family in cases where ED patients were isolated with family. Research is needed to confirm whether these differences were due to COVID-19 pandemic environmental influences or arise from the mode of treatment delivery.

Heterogeneity (I^2^) was a concern in the OP studies reducing confidence in the pooled OP findings. Publication bias was judged to be undetected and sensitivity analyses demonstrated the results to be robust across variation in study quality and type of ED; thus, increasing confidence in the pooled estimates.

#### GRADE Ratings

The strength of the whole body of evidence, the GRADE ratings for all the outcomes of the included studies were very low except for the DC-IOP studies for which the EDE-Q Global score was rated slightly better, as low. The GRADE ratings could be improved by reducing heterogeneity via the use of change-from-baseline between group effect sizes and by comparing equivalent treatment programmes with homogenous participants (e.g., matching severity and type of ED); increasing reporting and methodological quality; and utilising larger sample sizes to increase precision.

#### Comparison to the Literature

The extant literature indicates that online OP treatment is a promising alternative to in-person treatment. Outpatient cognitive behaviour therapy (CBT) and family therapy delivered online have shown comparable effectiveness to in-person delivery with similar therapeutic alliance and satisfaction with both modes of treatment delivery (Couturier et al., 2021). Additionally, treatment completion rate (adherence) has been found to be higher for online versus in-person treatment in outpatient mental health clinics prior to the pandemic (Mishkind et al., 2021).

There is less evidence for the effectiveness of DC-IOP, particularly in adult populations. Blalock et al. (2020) reported that an online IOP for adults was feasible, acceptable and improved ED symptoms, mood, self-esteem, and quality of life. Plumley et al. (2021) described a DC for adults which was transitioned to online delivery during the COVID-19 pandemic; improvements were found for BMI, ED symptoms and mood with high patient-rated therapeutic alliance. Online DC was compared to in-person DC treatment by Zimmerman et al. (2021) for a range of psychiatric disorders including eating disorders. Both modes of treatment delivery were comparable for patient satisfaction, reduced symptoms and suicidality, better functioning and coping ability, as well as positive mental health and general wellbeing. The only differences noted were that length of stay was longer (so possibly treatment response was slower) and treatment completion rate (adherence) was higher for the online cohort. Moreover, some patients commented that they would not have opted for in-person treatment even without pandemic restrictions, thus highlighting the appeal of online/mixed care.

In the UK Stewart et al. (2021) explored the transition to online OP ED treatment delivery mode early in the COVID-19 pandemic in a small sample of young people, carers and clinicians. Findings showed treatment was satisfactory, therapeutic alliance was adequate and technological competency was reported. Patients who transitioned from in-person to online care rather than starting care online during lockdown found therapy less effective. Patients and their carers differed on their preferences for treatment once the restrictions were lifted, with the latter being more likely to opt for online rather than in-person appointments. It is highlighted that the increased accessibility offered by online care especially for underserved populations needs to be balanced with an acceptable level of therapeutic engagement.

A study in the same service two years later reported that online OP ED treatment delivery mode may be acceptable even if not preferred by those seeking and delivering ED treatment. It was observed that attitudes toward online treatment post-pandemic remained mostly the same as during the pandemic despite both in-person and online modes now typically being offered in the UK beyond the forced switch implemented during the COVID-19 pandemic. Young people, carers and clinicians continued to prefer the initial assessment, early treatment sessions and the last session to be in-person, with the middle and latter sessions being slightly more preferred in a mixed delivery mode by carers in particular. In this study forming therapeutic alliance and talking about emotional issues was found to be easier via in-person mode (Baudinet et al., 2023).

However, as Stewart et al. (2021) and Baudinet et al. (2023) both used a sample of young people in an outpatient setting the findings while likely to be relevant for adult patients and DC-IOP treatment as well, need to be replicated in the latter population/treatment mode.

Evidence on the comparative effectiveness of in-person and online versus mixed (blended/hybrid) ED treatment delivery in the community is limited. In this systematic review and meta-analysis only one study compared all three treatment delivery modes reporting non-significant differences between the different modes, which is encouraging as it shows promise also for mixed mode ED community treatment delivery (Robinson et al., 2024). Ferrao Nunes-Zlotkowski et al. (2024) have confirmed in a systematic review and meta-analysis that mixed mode psychotherapy was feasible when in-person and online components were combined rather than sequential, and effectiveness was demonstrated for treating depression although findings for anxiety were non-significant. Cook-Cottone et al. (2024) and Novack and Chadi (2025) described mixed care as a promising mode of ED treatment delivery following the COVID-19 pandemic transition to online care, but the literature on this mode of treatment is still nascent.

Going forward the appropriate treatment delivery mode (in-person, online or mixed) may be best personalised to the individual, their family/carer and the clinician. Flexible change of mode may be indicated by treatment stage, treatment response/non-response, deterioration in mental/physical wellbeing, the need for in vivo (at home) support for new behaviours or when access is problematic.

#### Future Research

A notable limitation of online/mixed (blended/hybrid) ED treatment delivery is the risk to patients due to reduced accountability and difficulty in detecting and responding promptly to a crisis as options for immediate action are restricted. Moreover, the convenience of not leaving home for treatment and not having to be seen could promote avoidance and negatively impact the therapeutic alliance (Stewart et al., 2021). Patient safety and adverse effects of these treatments need to be investigated thoroughly, and systems put in place (e.g. in-person health monitoring where possible or daily check-ins with each patient) to support patients before online and mixed treatment delivery modes can be considered a viable option.

Telemedicine requires patients to have adequate literacy and IT skills, tends to reach a better educated, more affluent population and has been almost exclusively researched in White females (Wood et al., 2020). Despite benefits such as increased comfort, less disruption to education and family life, and reduced travel costs, online treatment places increased pressure on carers particularly in monitoring physical health risk at home (Serrano-Troncoso et al., 2020; Vuillier et al., 2021). Neurodiverse individuals, such as those with ADHD or autism, and those with social anxiety or who are physically vulnerable, may prefer online or mixed treatment; online treatment may attenuate pre-session anxiety and enhance ease of discussing sensitive issues (Baudinet et al., 2023; Webb et al., 2022). However, those with weak Internet connectivity, using their mobiles as their main device, and lacking private space which impacts on ease of discussing emotional difficulties (especially in domestic abuse situations) are likely to have a poorer therapeutic experience (Stewart et al., 2021).

It is necessary to address the requirements of those who are poorer, less educated, those with learning disability, severe co-morbid mental or medical illnesses, and males with EDs. In a US study Cachelin et al. (2000) reported that less acculturated ethnic minorities have a lower likelihood of seeking and receiving treatment for an ED and this warrants further research.

More recently, James et al. (2017) noted the need to ensure research using novel technologies adequately represents minority populations to reduce current health disparities. In four of the included studies (Blalock et al., 2025; Levinson et al., 2021; Ortiz et al., 2023; Penwell et al., 2023) despite the majority of patients being White there was a trend of greater representation of racial/ethnic minorities in the online group versus the in-person group; this preference requires more exploration.

Research is needed regarding patient and health provider barriers which hinder uptake of ED community care including stigma, denying the illness, low motivation to seek treatment, high cost of treatment, time constraints, lack of adequate IT skills, low mental health literacy, limited availability of trained therapists, and lack of local ED services in remote rural areas (Ali et al., 2017). In particular socioeconomic status, race - minority bias and literacy are factors that could impact engagement with online treatment, and more research is needed to attenuate these health inequities (James et al., 2017).

Older age, and low level of education indicative of low socio-economic status, are commonly cited barriers to uptake of telemedicine (Barbosa et al., 2021). Niemeyer and Knaevelsrud (2023) observed that individuals with low socio-economic status were less likely to be offered outpatient psychotherapy in Germany than those with moderate-high socio-economic status. However, contrary to reports that older adults may find it harder to engage with technological interventions Rienecke et al. (2024) observed high satisfaction with an online ED intensive outpatient programme (IOP) and no statistically significant differences were found between different age groups including adults aged over 40 years, albeit in a primarily female and White patient group. In Blalock et al. (2025) although the online IOP group consisted of significantly older patients than the in-person IOP group, both modes of treatment delivery were similarly effective suggesting that older age is not necessarily a barrier to engaging in online IOP treatment.

Other barriers to effective ED treatment include difficulties during transitions (e.g. from young people ED services to adult ED services; geographical - home to university services; and between inpatient and community - outpatient care), and inadequate availability of outpatient care provision leading to pressure being placed on primary care general practitioners (GPs) (Shekar, 2022); online care could be advantageous in such circumstances.

As well as research on improving the online therapeutic alliance, qualitative research is required to determine patient-carer-health provider values, preferences, satisfaction, and experiences regarding the different treatment delivery modes; and in particular to explore reasons for treatment dropout or non-improvement, in order to improve ED community care and maximise compliance. In addition, how treatment outcomes are affected by baseline severity of ED symptoms, type of ED diagnosis and demographics such as age, gender and ethnicity need to be investigated to optimise ED community care. It is necessary to understand who would or would not benefit from the different modes of treatment delivery to provide targeted individualised patient-centred care and to determine whether online/mixed treatment delivery modes do increase access to evidence-based treatment for underserved populations.

Better consistent reporting of participant retention, follow-up data, adverse effects, co-morbidities, co-interventions, history of ED, any past ED treatment, inclusion of minority groups and particularly reporting of socioeconomic status is recommended. Protective factors need further exploration; improved self-esteem, coping mechanisms and quality of life. It is important to identify the specific components of in-person, online and mixed delivery modes that are most effective for the community treatment of EDs to provide an individualised and optimal dose of care. Research is essential to clarify whether the findings can be transferred to countries beyond high income western countries and to different healthcare systems. Further, cost-effectiveness trials are required to support the argument for the widespread implementation of online/mixed modes of ED community care delivery with regard to resource use and opportunity cost; mixed delivery mode in particular could reduce costs and retain effectiveness.

Meal support, group and family/multi-family sessions are particularly difficult to implement online and require more development (Baudinet et al., 2023; Webb et al., 2022). Brothwood et al. (2021) observed that between therapy social interactions that occur in in-person treatment are lost in online treatment and alternatives need to be found to create deeper connections; trusting relationships online. Beat (2024) noted that peer relationships, inter-carer connections and therapeutic alliance were difficult to develop in online treatment and proposed that mixed mode treatment may attenuate this challenge by, for example, offering initial in-person sessions; more research on mixed care is recommended.

Matthews et al. (2019) suggested that patients should be motivationally ready to engage with intensive ED treatment programmes such as daycare (DC). It has been observed that patients with AN were the least satisfied with online therapy compared to patients with other ED diagnoses (Fernandez-Aranda, Munguia, et al., 2020); this may be due to the ego-syntonic nature of their illness and high ambivalence towards recovery which may impede engagement with treatment (Abbate-Daga et al., 2013). Those with AN may benefit from increased support, the nature of which should be a focus of future research.

Long-term outcomes of online and mixed treatment delivery need to be evaluated in relation to in-person care. Baudinet et al. (2023) reported that current evidence indicates therapeutic alliance and early treatment response significantly affect post-treatment outcomes and as the provision of online treatment early in treatment may prolong treatment it is important for future research to determine whether this leads to worse outcomes.

Therapists can be less satisfied with online/mixed (blended/hybrid) modes of psychotherapy than patients and carers; this barrier to telemedicine implementation needs to be addressed to maximise treatment benefits (Shaw et al., 2021). McFarlane et al. (2015) suggested implementing tailored approaches encompassing patient preferences and Zimmerman et al. (2021) highlighted that compensation (insurance reimbursement) for online/mixed treatment modes needs to be in parity with in-person treatment delivery to ensure virtual care is available as a viable treatment option.

Particularly relevant, Catenacci and Couturier (2023) have suggested specific improvements to online daycare (DC) programmes that merit exploration: (1) increasing involvement and support for family/carers; (2) screening suitability for online care; (3) providing more meal support and increased frequency of in-person check-ins; (4) managing rules and patient expectations in advance; (5) increasing communication with healthcare providers; (6) ensuring patients have a reliable Internet connection; and (7) utilising a hybrid delivery mode.

Finally, the field would benefit from more high-quality quasi-experimental (QE) studies allowing for change score meta-analyses and ideally randomised controlled trials (RCTs) with adequate power are needed so that effect sizes can be definitively determined. Before dissemination and implementation positive results would require replication outside of pandemic times and in large high-quality RCTs with lengthy follow-up.

### Limitations

The main drawback of this systematic review and meta-analysis is that only an independent researcher conducted the review processes, however, this was under PhD supervision. Other drawbacks include: (1) the OP studies did not provide the data required to conduct change score meta-analyses so the less reliable post-treatment between group effect sizes (without change scores) were calculated; (2) grey literature was not searched but it is contended that the search was exhaustive; (3) all online samples were studied during the pandemic, thus environmental issues may have affected the results; 4) the outcome measures meta-analysed were all patient reported outcome measures (PROMs) which are liable to recall bias; and (5) certainty in the findings of this review is attenuated by the small number of included studies and only one study compared mixed (blended/hybrid) treatment mode to online and in-person modes (Robinson et al., 2024). However, in the absence of ‘gold standard’ randomised controlled trials (RCTs), the quasi-experimental (QE) studies included in this systematic review and meta-analysis provide the best evidence base available at this time.

### Conclusions

Eating disorder (ED) telemedicine treatment programmes could reduce the financial burden to the healthcare system, patients, their families, and facilitate earlier intervention mitigating the disruption to social functioning, education and employment commitments/prospects of ED sufferers, improving their overall quality of life. Online and mixed (blended/hybrid) delivery modes of ED community care could increase access to evidence-based treatment and reduce health disparities associated with, for example, socio-economic status, race/ethnicity, gender and sexual minority groups, geographic location (rural, remote regions) and disability. This systematic review and meta-analysis provides preliminary support for the comparative effectiveness, safety and acceptability of ED community care provision in adult services via in-person, online or mixed delivery modes in western high-income countries. It is proposed that telemedicine, in particular mixed delivery mode, although not a replacement could complement existing in-person ED services. However, the evidence base is very small, particularly for the outpatient studies and mixed treatment delivery mode. Further research is necessary using rigorous high-quality adequately powered quasi-experimental (QE) and ideally randomised controlled trial (RCT) designs to elucidate the effectiveness and efficacy of treatment outside of the COVID-19 pandemic context. In a rapidly evolving field, for a highly vulnerable population, it is imperative to exploit new technologies to optimise the delivery of ED treatment, with the goal of providing timely, flexible, personalised, cost-effective, and evidence-based interventions.

## Ethical Considerations

This work was secondary research and did not involve the use of human subjects thus there were no ethical considerations and lived experience contributors were not involved.

## Data Availability Statement

The data that support the findings of this study are available from the corresponding author upon reasonable request.

## Acknowledgements

This journal article is part of the thesis submission for the award of PhD in Medicine (University College London, UK) by Nilima Hamid.

## Author Contributions

The co-author contributed to this article as a PhD primary academic supervisor.

## REFERENCES

Abbate-Daga, G., Amianto, F., Delsedime, N., De-Bacco, C., & Fassino, S. (2013). Resistance to treatment in eating disorders: A critical challenge. BMC Psychiatry 13:294.

Abbate-Daga, G., Marzola, E., De-Bacco, C., Buzzichelli, S., Brustolin, A., Campisi, S., Amianto, F., Migliaretti, G., & Fassino, S. (2015). Day hospital treatment for Anorexia Nervosa: A 12-month follow-up study. Eur Eat Disord Rev, 23(5), 390–398. 10.1002/erv.2369

Ali, K., Farrer, L., Fassnacht, D. B., Gulliver, A., Bauer, S., & Griffiths, K. M. (2017). Perceived barriers and facilitators towards help-seeking for eating disorders: A systematic review. International Journal of Eating Disorders, 50(1), 9–21. 10.1002/eat.22598

Barbosa, W., Zhou, K., Waddell, E., Myers, T., & Dorsey, E. R. (2021). Improving Access to Care: Telemedicine Across Medical Domains. Annual Review of Public Health, 42, 463–481. 10.1146/annurev-publhealth-090519-093711

Barker, T. H., Habibi, N., Aromataris, E., Stone, J. C., Leonardi-Bee, J., Sears, K., Hasanoff, S., Klugar, M., Tufanaru, C., Moola, S., & Munn, Z. (2024). The revised JBI critical appraisal tool for the assessment of risk of bias for quasi-experimental studies. JBI Evidence Synthesis, 22(3), 378–388. 10.11124/JBIES-23-00268

Barrett, E., & Richardson, S. C. (2020). Eating disorders during the COVID-19 pandemic. Irish Medical Journal, 114(1).

Baudinet, J., Konstantellou, A., Hambleton, A., Bialluch, K., Hurford, G., & Stewart, C. S. (2023). Do people want the ‘new normal’? A mixed method investigation of young person, parent, and clinician experience and preferences for eating disorder treatment delivery in the post-COVID-19 world [Article]. Nutrients, 15(17), Article 3732. 10.3390/nu15173732

Beat. (2024). There’s no place like home: The case for intensive community and day treatments for eating disorders.

Beck, A. T., Steer, R. A., & Garbin, M. G. (1988). Psychometric properties of the Beck Depression Inventory: Twenty-five years of evaluation. Clinical Psychology Review, 8(1), 77–100.

Blalock, D. V., Le Grange, D., Johnson, C., Duffy, A., Manwaring, J., Tallent, C. N., Schneller, K., Solomon, A. M., Mehler, P. S., McClanahan, S. F., & Rienecke, R. D. (2020). Pilot assessment of a virtual intensive outpatient program for adults with eating disorders. European Eating Disorders Review, 28(6), 789–795. 10.1002/erv.2785

Blalock, D. V., Mehler, P. S., Michel, D. M., Duffy, A., Le Grange, D., O’Melia, A. M., & Rienecke, R. D. (2025). Virtual versus in-person intensive outpatient treatment for eating disorders during the COVID-19 pandemic in United States-based treatment facilities: Naturalistic study [Comparative Study]. Journal of Medical Internet Research, 27, e66465. 10.2196/66465

Brothwood, P. L., Baudinet, J., Stewart, C. S., & Simic, M. (2021). Moving online: young people and parents’ experiences of adolescent eating disorder day programme treatment during the COVID-19 pandemic. Journal of Eating Disorders, 9(1), Article 62. 10.1186/s40337-021-00418-4

Cachelin, F. M., Veisel, C., Barzegarnazari, E., & Striegel-Moore, R. H. (2000). Disordered eating, acculturation, and treatment-seeking in a community sample of Hispanic, Asian, Black, and White Women. Psychology of Women Quarterly, 24, 244–253.

Campbell, S., Maunder, K., Lehmann, O., McKeown, M., & McNicholas, F. (2022). The long-term impact of COVID-19 on presentations to a specialist child and adolescent eating disorder program. Irish Medical Journal, 115(8).

Carr, A., Toloza, C., Li, Z., Nazar, B. P., & Himmerich, H. (2022). Therapy outcome of day treatment for people with anorexia nervosa before and during the COVID-19 pandemic. Brain and Behavior, 12(6), Article e2604. 10.1002/brb3.2604

Catenacci, V., & Couturier, J. (2023). Experiences of a virtual day program for adolescents with eating disorders: A qualitative analysis of benefits and barriers. Journal of Eating Disorders, 11(1), Article 133. 10.1186/s40337-023-00859-z

Cohen, J. (1988). Statistical power analysis for the behavioral sciences (2nd ed.). Lawrence Erlbaum Associates

Cook-Cottone, C., Harriger, J. A., Tylka, T. L., & Wood-Barcalow, N. L. (2024). Virtually possible: strategies for using telehealth in eating disorder treatment learned from the COVID-19 pandemic. Eating Disorders, 32(2), 99–119. 10.1080/10640266.2023.2261762

Couturier, J., Pellegrini, D., Miller, C., Bhatnagar, N., Boachie, A., Bourret, K., Brouwers, M., Coelho, J. S., Dimitropoulos, G., Findlay, S., Ford, C., Geller, J., Grewal, S., Gusella, J., Isserlin, L., Jericho, M., Johnson, N., Katzman, D. K., Kimber, M.,…Webb, C. (2021). The COVID-19 pandemic and eating disorders in children, adolescents, and emerging adults: Virtual care recommendations from the Canadian consensus panel during COVID-19 and beyond. Journal of Eating Disorders, 9(1), Article 46. 10.1186/s40337-021-00394-9

Devoe, D. J., Han, A., Anderson, A., Katzman, D. K., Patten, S. B., Soumbasis, A., Flanagan, J., Paslakis, G., Vyver, E., Marcoux, G., & Dimitropoulos, G. (2023). The impact of the COVID-19 pandemic on eating disorders: A systematic review. International Journal of Eating Disorders, 56(1), 5–25. 10.1002/eat.23704

Fairburn, C. G., & Beglin, S. J. (1994). Assessment of eating disorders: Interview or self-report questionnaire? International Journal of Eating Disorders, 16(4), 363–370. 10.1002/1098-108x(199412)16:4<363::Aid-eat2260160405>3.0.Co;2-#

Fairburn, C. G., & Patel, V. (2014). The global dissemination of psychological treatments: A road map for research and practice. American Journal of Psychiatry, 171(5), 495–498.

Feldhege, J., Moessner, M., Wolf, M., & Bauer, S. (2021). Changes in language style and topics in an online eating disorder community at the beginning of the COVID-19 pandemic: Observational study. Journal of Medical Internet Research, 23(7), e28346. 10.2196/28346

Fernandez-Aranda, F., Casas, M., Claes, L., Bryan, D. C., Favaro, A., Granero, R., Gudiol, C., Jimenez-Murcia, S., Karwautz, A., Le Grange, D., Menchon, J. M., Tchanturia, K., & Treasure, J. (2020). COVID-19 and implications for eating disorders. European Eating Disorders Review, 28(3), 239–245. 10.1002/erv.2738

Fernandez-Aranda, F., Munguia, L., Mestre-Bach, G., Steward, T., Etxandi, M., Baenas, I., Granero, R., Sanchez, I., Ortega, E., Andreu, A., Moize, V. L., Fernandez-Real, J. M., Tinahones, F. J., Dieguez, C., Fruhbeck, G., Le Grange, D., Tchanturia, K., Karwautz, A., Zeiler, M.,…Jimenez-Murcia, S. (2020). COVID Isolation Eating Scale (CIES): Analysis of the impact of confinement in eating disorders and obesity-A collaborative international study. European Eating Disorders Review, 28(6), 871–883. 10.1002/erv.2784

Ferrao Nunes-Zlotkowski, K., Shepherd, H. L., Beatty, L., Butow, P., & Shaw, J. M. (2024). Blended psychological therapy for the treatment of psychological disorders in adult patients: Systematic review and meta-analysis. Interactive Journal of Medical Research, 13, e49660. 10.2196/49660

Friars, D., Walsh, O., & McNicholas, F. (2023). Assessment and management of cardiovascular complications in eating disorders. Journal of Eating Disorders, 11(1), 13. 10.1186/s40337-022-00724-5

Gorrell, S., Reilly, E. E., Brosof, L., & Le Grange, D. (2022). Use of telehealth in the management of adolescent eating disorders: Patient perspectives and future directions suggested from the COVID-19 pandemic. Adolescent Health, Medicine and Therapeutics, 13, 45–53. 10.2147/AHMT.S334977

Hamid, N. (2023). Internet-based cognitive behaviour therapy for the prevention, treatment and relapse prevention of eating disorders: A systematic review and meta-analysis. Psych Journal, 13(1), 5–18. 10.1002/pchj.715

Higgins, J. P. T., & Thomas, J. (2019). Cochrane handbook of systematic reviews of interventions (2nd ed.). John Wiley & Sons.

Irish, M., Dalton, B., Potts, L., McCombie, C., Shearer, J., Au, K., Kern, N., Clark-Stone, S., Connan, F., Johnston, A. L., Lazarova, S., Macdonald, S., Newell, C., Pathan, T., Wales, J., Cashmore, R., Marshall, S., Arcelus, J., Robinson, P.,…Schmidt, U. (2022). The clinical effectiveness and cost-effectiveness of a ‘stepping into day treatment’ approach versus inpatient treatment as usual for anorexia nervosa in adult specialist eating disorder services (DAISIES trial): A study protocol of a randomised controlled multi-centre open-label parallel group non-inferiority trial. Trials [Electronic Resource], 23(1), 500. 10.1186/s13063-022-06386-7

James, D. C., Harville, C., 2nd, Sears, C., Efunbumi, O., & Bondoc, I. (2017). Participation of African Americans in e-health and m-health studies: A systematic review. Telemedicine Journal and e-Health, 23(5), 351–364. 10.1089/tmj.2016.0067

Kroenke, K., Spitzer, R. L., & Wiliams, J. B. (2001). The PHQ-9: Validity of a brief depression severity measure. Journal of General Internal Medicine, 16(9), 606–613.

Levinson, C. A., Spoor, S. P., Keshishian, A. C., & Pruitt, A. (2021). Pilot outcomes from a multidisciplinary telehealth versus in-person intensive outpatient program for eating disorders during versus before the Covid-19 pandemic [Empirical Study; Quantitative Study]. International Journal of Eating Disorders, 54(9), 1672–1679. 10.1002/eat.23579

Matthews, K., Gordon, L., van Beusekom, J., Sheffield, J., & Patterson, S. (2019). A day treatment program for adults with eating disorders: Staff and patient experiences in implementation. Journal of Eating Disorders, 7, Article 21. 10.1186/s40337-019-0252-4

McFarlane, T., MacDonald, D. E., Trottier, K., & Olmsted, M. P. (2015). The effectiveness of an individualized form of day hospital treatment. Eating Disorders, 23(3), 191–205. 10.1080/10640266.2014.981430

Mishkind, M. C., Shore, J. H., Bishop, K., D’Amato, K., Brame, A., Thomas, M., & Schneck, C. D. (2021). Rapid conversion to telemental health services in response to COVID-19: Experiences of two outpatient mental health clinics. Telemedicine Journal and e-Health, 27(7), 778–784. 10.1089/tmj.2020.0304

Moher, D., Liberati, A., Tetzlaff, J., Altman, D. G., & Group, P. (2009). Preferred reporting items for systematic reviews and meta-analyses: The PRISMA statement. PLoS Medicine, 6(7), e1000097. 10.1371/journal.pmed.1000097

Nelson, H. D. (2025). Systematic reviews to answer health care questions (2nd ed.). Wolters Kluwer.

Niemeyer, H., & Knaevelsrud, C. (2023). Socioeconomic status and access to psychotherapy. Journal of Clinical Psychology, 79(4), 937–953. 10.1002/jclp.23449

Norman, G. R., Sloan, J. A., & Wyrwich, K. W. (2003). Interpretation of changes in health-related quality of life. The remarkable universality of half a standard deviation. Medical Care, 41(5), 582–592.

Norman, S. B., Cissell, S. H., Means-Christensen, A. J., & Stein, M. B. (2006). Development and validation of an Overall Anxiety Severity and Impairment Scale (OASIS). Depression and Anxiety, 23(4), 245–249.

Novack, K., & Chadi, N. (2025). The hybrid space in eating disorder treatment: Towards a personalized approach to integrating telehealth and in-person care. Journal of Eating Disorders, 13(1), 22. 10.1186/s40337-025-01211-3

Obiekezie, A., Friel, C., & Pathan, M. T. (2022). Safety of delivering eating disorders day treatment programme on the virtual platform in (COVID-19) pandemic [Conference Abstract]. BJPsych Open, 8(Supplement 1), S8. 10.1192/bjo.2022.91

Orme, J. G., Reis, J., & Herz, E. J. (1986). Factorial and discriminant validity of the Center for Epidemiological Studies Depression (CES-D)scale. Journal of Clinical Psychology, 42(1), 28–33.

Ortiz, A. M. L., Cusack, C. E., Billman, M. G., & Essayli, J. H. (2023). Baseline symptomatology and treatment outcomes of young adults in a virtual versus in-person partial hospitalization and intensive outpatient program for eating disorders [Empirical Study; Quantitative Study; Treatment Outcome]. International Journal of Eating Disorders, 56(8), 1644–1649. 10.1002/eat.23968

Page, M. J., McKenzie, J. E., Bossuyt, P. M., Boutron, I., Hoffmann, T. C., Mulrow, C. D., Shamseer, L., Tetzlaff, J. M., Akl, E. A., Brennan, S. E., Chou, R., Glanville, J., Grimshaw, J. M., Hrobjartsson, A., Lalu, M. M., Li, T., Loder, E. W., Mayo-Wilson, E., McDonald, S.,…Moher, D. (2021). The PRISMA 2020 statement: An updated guideline for reporting systematic reviews. BMJ, 372, n71. 10.1136/bmj.n71

Penwell, T. E., Smith, M., Ortiz, S. N., Brooks, G., & Thompson-Brenner, H. (2023). Traditional versus virtual partial hospital programme for eating disorders: Feasibility and preliminary comparison of effects [Empirical Study; Quantitative Study]. European Eating Disorders Review, 32(2), 163–178. 10.1002/erv.3031

Pilkonis, P. A., Choi, S. W., Reise, S. P., Stover, A. M., Riley, W. T., & Cella, D. (2011). Item banks for measuring emotional distress from the Patient-Reported Outcomes Measurement Information System (PROMIS): Depression, anxiety, and anger. Assessment, 18(3), 263–283.

Plumley, S., Kristensen, A., & Jenkins, P. E. (2021). Continuation of an eating disorders day programme during the COVID-19 pandemic [Empirical Study; Quantitative Study]. Journal of Eating Disorders Vol 9 2021, ArtID 34, 9. 10.1186/s40337-021-00390-z

Raykos, B. C., Erceg-Hurn, D. M., Hill, J., Campbell, B. N., & McEvoy, P. M. (2021). Positive outcomes from integrating telehealth into routine clinical practice for eating disorders during COVID-19 [Empirical Study; Interview; Quantitative Study]. International Journal of Eating Disorders, 54(9), 1689–1695. 10.1002/eat.23574

Rienecke, R. D., Blalock, D. V., Tallent, C. N., Duffy, A., & Mehler, P. S. (2024). Eating disorder virtual intensive outpatient program: Patient satisfaction according to age group. Eating Disorders, 32(5), 493–508. 10.1080/10640266.2024.2325296

Rivera, V., Aldridge, M. D., Ornstein, K., Moody, K. A., & Chun, A. (2021). Racial and socioeconomic disparities in access to telehealth. Journal of the American Geriatrics Society, 69(1), 44–45. 10.1111/jgs.16904

Robinson, P., Cauvi, S., Morando, S., & Beckford, L. (2024). In malnourished patients with anorexia nervosa online day care appears to be as effective as in person [Conference Abstract]. Clinical Nutrition ESPEN, Conference: 46th ESPEN Congress on Clinical Nutrition & Metabolism. Milan Italy. 63, 1316. 10.1016/j.clnesp.2024.07.1035

Schlissel, A. C., Richmond, T. K., Eliasziw, M., Leonberg, K., & Skeer, M. R. (2023). Anorexia nervosa and the COVID-19 pandemic among young people: A scoping review. Journal of Eating Disorders, 11(1), Article 122. 10.1186/s40337-023-00843-7

Serrano-Troncoso, E., Fàbrega-Ribera, M., Coll-Pla, N., Godrid-García, M., Carulla-Roig, M., Cecilia-Costa, R., Soto-Usera, P., Sánchez-Fernández, B., Matalí-Costa, J., & Dolz-Abadia, M. (2020). Alternatives to inpatient treatment in adolescents with anorexia nervosa: Effectiveness and characteristics of a new intensive model of day patient treatment. Actas Espanolas de Psiquiatria, 48(1), 19–27.

Shaw, H., Robertson, S., & Ranceva, N. (2021). What was the impact of a global pandemic (COVID-19) lockdown period on experiences within an eating disorder service? A service evaluation of the views of patients, parents/carers and staff. Journal of Eating Disorders, 9(1), 14. 10.1186/s40337-021-00368-x

Shekar, D. (2022). Barriers to eating disorder treatment: A review of socioeconomic obstacles and gaps in the healthcare system. Research Archive of Rising Scholars.

Silen, Y., & Keski-Rahkonen, A. (2022). Worldwide prevalence of DSM-5 eating disorders among young people. Current Opinion in Psychiatry, 35(6), 362–371. 10.1097/YCO.0000000000000818

Spitzer, R. L., Kroenke, K., Williams, J. B. W., & Lowe, B. (2006). A brief measure for assessing generalized anxiety disorder: The GAD-7. Archives of Internal Medicine, 166(10), 1092–1097.

Steiger, H., Booij, L., Crescenzi, O., Oliverio, S., Singer, I., Thaler, L., St-Hilaire, A., & Israel, M. (2021). In-person versus virtual therapy in outpatient eating-disorder treatment: A COVID-19 inspired study [Empirical Study; Quantitative Study]. International Journal of Eating Disorders, 55(1), 145–150. 10.1002/eat.23655

Stewart, C., Konstantellou, A., Kassamali, F., McLaughlin, N., Cutinha, D., Bryant-Waugh, R., Simic, M., Eisler, I., & Baudinet, J. (2021). Is this the ‘new normal’? A mixed method investigation of young person, parent and clinician experience of online eating disorder treatment during the COVID-19 pandemic. Journal of Eating Disorders, 9(1), 78. 10.1186/s40337-021-00429-1

Thaler, L., Booij, L., St-Hilaire, A., Paquin-Hodge, C., Mesli, N., Burko, H., Lee, V., Oliverio, S., Israel, M., & Steiger, H. (2024). Outcomes of a virtual day treatment program for adults with eating disorders-comparison with in-person day treatment [Empirical Study; Quantitative Study]. International Journal of Eating Disorders, No Pagination Specified. 10.1002/eat.24263

Vuillier, L., May, L., Greville-Harris, M., Surman, R., & Moseley, R. L. (2021). The impact of the COVID-19 pandemic on individuals with eating disorders: The role of emotion regulation and exploration of online treatment experiences. Journal of Eating Disorders, 9(1), 10. 10.1186/s40337-020-00362-9

Webb, H., Dalton, B., Irish, M., Mercado, D., McCombie, C., Peachey, G., Arcelus, J., Au, K., Himmerich, H., Johnston, A. L., Lazarova, S., Pathan, T., Robinson, P., Treasure, J., Schmidt, U., & Lawrence, V. (2022). Clinicians’ perspectives on supporting individuals with severe anorexia nervosa in specialist eating disorder intensive treatment settings during the COVID-19 pandemic. Journal of Eating Disorders, 10(1), 30. 10.1186/s40337-022-00555-4

Wood, S. M., White, K., Peebles, R., Pickel, J., Alausa, M., Mehringer, J., & Dowshen, N. (2020). Outcomes of a rapid adolescent telehealth scale-up during the COVID-19 pandemic. Journal of Adolescent Health, 67(2), 172–178. 10.1016/j.jadohealth.2020.05.025

Zimmerman, M., Terrill, D., D’Avanzato, C., & Tirpak, J. W. (2021). Telehealth treatment of patients in an intensive acute care psychiatric setting during the COVID-19 pandemic: Comparative safety and effectiveness to in-person treatment [Article]. Journal of Clinical Psychiatry, 82(2), Article 20m13815. 10.4088/JCP.20M13815

Zipfel, S., Reas, D. L., Thornton, C., Olmsted, M. P., Williamson, D. A., Gerlinghoff, M., Herzog, W., & Beumont, P. J. (2002). Day hospitalization programs for eating disorders: A systematic review of the literature. International Journal of Eating Disorders, 31(2), 105–117. 10.1002/eat.10009

